# Enhanced CRISPR/Cas-Based Immunoassay through Magnetic Proximity Extension and Detection

**DOI:** 10.1101/2024.09.06.24313206

**Authors:** Fangchi Shao, Jiumei Hu, Pengfei Zhang, Patarajarin Akarapipad, Joon Soo Park, Hanran Lei, Kuangwen Hsieh, Tza-Huei Wang

**Author notes:** Department of Chemical and Biological Engineering, Princeton University, Princeton, New Jersey, 08544, United States (Pengfei Zhang). Equal contribution between Fangchi Shao and Jiumei Hu. Co-corresponding authors &.

## Abstract

Clustered regularly interspaced short palindromic repeats (CRISPR)/Cas-associated systems have recently emerged as a focal point for developing next-generation molecular diagnosis, particularly for nucleic acid detection. However, the detection of proteins is equally critical across diverse applications in biology, medicine, and the food industry, especially for diagnosing and prognosing diseases like cancer, Alzheimer’s and cardiovascular conditions. Despite recent efforts to adapt CRISPR/Cas systems for protein detection with immunoassays, these methods typically achieved sensitivity only in the femtomolar to picomolar range, underscoring the need for enhanced detection capabilities. To address this, we developed CRISPR-AMPED, an innovative CRISPR/Cas-based immunoassay enhanced by magnetic proximity extension and detection. This approach combines proximity extension assay (PEA) with magnetic beads that converts protein into DNA barcodes for quantification with effective washing steps to minimize non-specific binding and hybridization, therefore reducing background noise and increasing detection sensitivity. The resulting DNA barcodes are then detected through isothermal nucleic acid amplification testing (NAAT) using recombinase polymerase amplification (RPA) coupled with the CRISPR/Cas12a system, replacing the traditional PCR. This integration eliminates the need for thermocycling and bulky equipment, reduces amplification time, and provides simultaneous target and signal amplification, thereby significantly boosting detection sensitivity. CRISPR-AMPED achieves attomolar level sensitivity, surpassing ELISA by over three orders of magnitude and outperforming existing CRISPR/Cas-based detection systems. Additionally, our smartphone-based detection device demonstrates potential for point-of-care applications, and the digital format extends dynamic range and enhances quantitation precision. We believe CRISPR-AMPED represents a significant advancement in the field of protein detection.

## Introduction

Clustered regularly interspaced short palindromic repeats (CRISPR)/CRISPR-associated systems (Cas) have in recent years emerged as the center of interest for developing next-generation molecular diagnostics^1, 2^. While CRISPR/Cas-based detection of nucleic acids has garnered the most attention and research efforts thus far^3-6^, detection of proteins is also important for a wide ranging applications in biology, medicine, and food industry^7-13^, and especially for the diagnosis and prognosis of various diseases, including cancers^14-16^, Alzheimer’s diseases^9, 17, 18^, and cardiovascular diseases^19, 20^. To date, there has seen a number of publications using CRISPR for protein detection^21-23^. Furthermore, the applications of CRISPR have been extended to the detection of proteins when coupled with immunosorbent assays. However, the sensitivity of such approaches has generally been limited to the femtomolar to picomolar level^24-27^. Thus, there remains a need to further advance CRISPR/Cas-based protein detection.

Proximity extension assay (PEA) is an emerging technique that couples antibody-based binding and DNA-enabled amplification for sensitive and specific protein detection^28-32^. More specifically, PEA utilizes a pair of antibodies that are conjugated with DNA probes containing a short complementary region such that binding between the protein target and the antibody pair stabilizes the hybridization between DNA probes, enables subsequent DNA extension, and facilitates downstream PCR-based amplification and detection. More recently, PEA has been adapted into the magnetic format by incorporating another magnetically-labeled antibody^33^. The addition of the magnetically-labeled antibody opens the door to washing steps that can reduce nonspecific binding and nonspecific hybridization, thereby improving the detection sensitivity. The advent of magnetic PEA offers a promising route for augmenting CRISPR/Cas-based protein detection methods. To date, however, such a combination has yet to be explored.

In response, we developed CRISPR/Cas-based immunoassay augmented by magnetic proximity extension and detection (CRISPR-AMPED) and demonstrated that this combination indeed led to highly sensitive protein detection and an array of additional benefits. To do so, we drew inspiration from nucleic acid detection methods^34-38^ and established a rapid one-step reaction with concurrent isothermal DNA amplification and CRISPR/Cas-based fluorescence detection for further sensitivity enhancement without sacrificing assay turnaround time. CRISPR-AMPED could achieve attomolar detection, which outpaced ELISA by > 3 orders of magnitude. Amplified fluorescence signal from CRISPR-AMPED not only could be detected by standard benchtop fluorescence detector but also a custom smartphone attachment imaging module, thus opening the door to portable detection and enhancing point-of-care (POC) amenability. Additionally, CRISPR-AMPED could be adapted in the digital format, which expanded detection dynamic range and improved quantitation precision. Taken together, we demonstrated CRISPR-AMPED as a potentially powerful method for protein detection toward a wide range of applications.

## Results and Discussion

### Overview of CRISPR-AMPED

CRISPR-AMPED commences with the magnetic proximity extension step (Figure 1A). During this step, the protein target is initially bound with the magnetically-labeled capture antibody and subsequently bound with the pair of DNA probe-labeled detection antibodies (i.e., PEA probes). The protein-antibody complex stabilizes the hybridization between DNA probes that share a short 5-bp complementary region. Here, the use of magnetic beads enables extensive washing, reducing potential background noise and therefore boosting the assay sensitivity. Moreover, the use of triple-antibody binding – one from magnetically-labeled capture antibody and two from DNA probe-labeled detection antibody pair – can also bolster assay specificity (Figure S1). The 5-bp hybridized region between the DNA probes then serves as the site for sequence extension using *Bst* polymerase, which results in a double-stranded DNA extension product that can serve as the trigger for downstream reaction.

**Figure 1:**
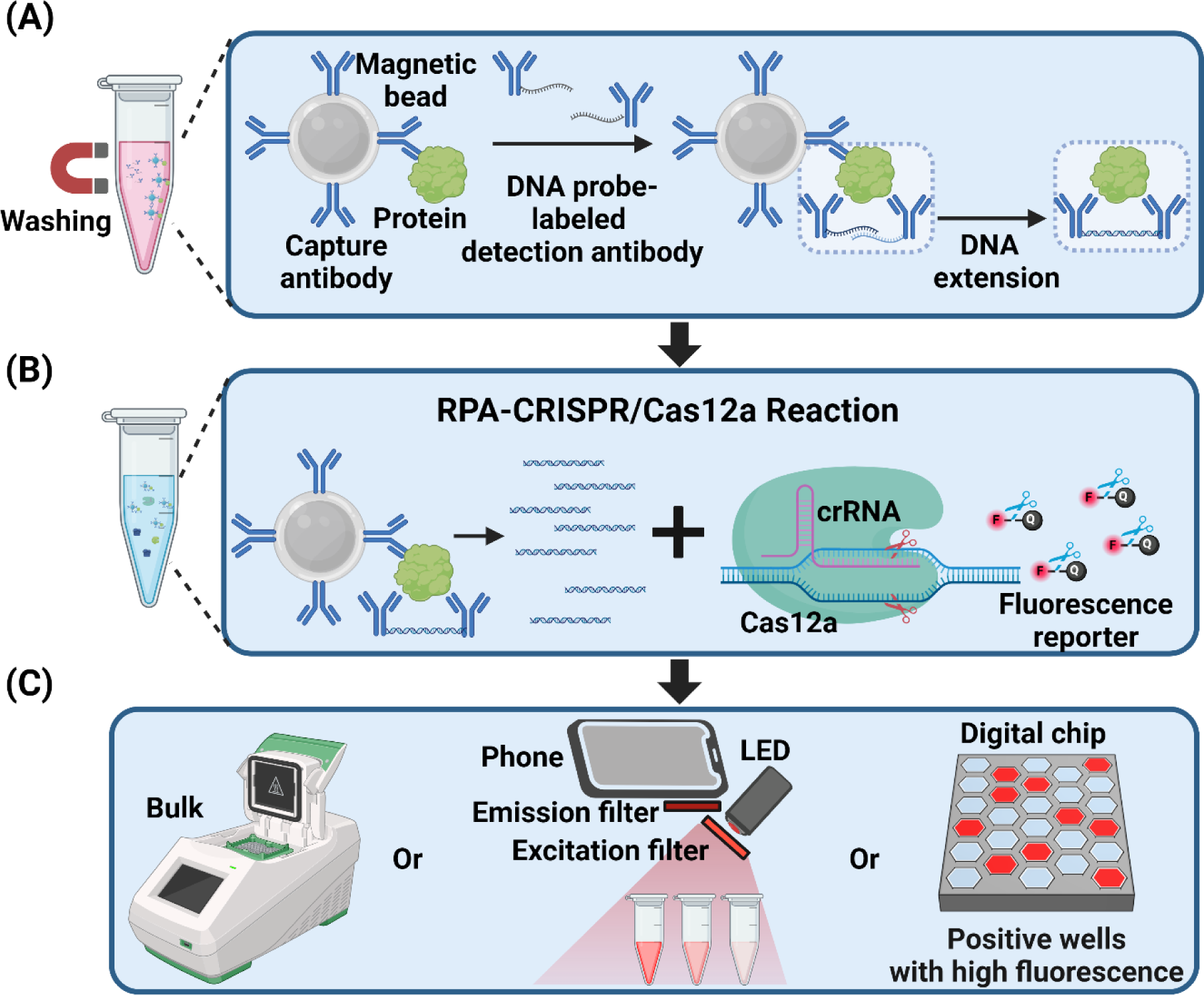
CRISPR-AMPED. (A) CRISPR-AMPED begins with the capture of protein biomarker with magnetically-labeled antibody. A pair of detection antibodies labeled with DNA probe (i.e., PEA probe) are then added to bind to the protein, where the DNA probes in proximity will be extended to double stranded DNA sequence with the aid of *Bst* polymerase. All washing steps are enabled by magnetic beads to remove unbound proteins and PEA probes. (B) The extended DNA sequence is amplified by RPA and detected with CRISPR/Cas12a-cleavge of fluorescence reporters to yield fluorescence signal. (C) The assay has been demonstrated with three downstream detection platforms, bulk Bio-Rad CFX real-time PCR detection system, smartphone-based detection device, and digital chips.

The downstream reaction combines recombinase polymerase amplification (RPA) and CRISPR/Cas12a-based reaction in a single step (Figure 1B). Here, the combination of RPA and CRISPR/Cas12a not only has shown the capacity for sensitive nucleic acid detection^34-36, 38-42^, but is also commercially available and thus easily accessible. For this single-step reaction, we adopted our previously developed strategy^34^ using a fine-tuned ratio between RPA primers and the complexes of CRISPR guide RNA (crRNA) and Cas12a effector such that RPA could amplify the double-stranded DNA extension product into numerous copies of DNA activators for Cas12a effector while activated Cas12a effector could concurrently cleave single-stranded DNA fluorescence reporters, thus producing strong fluorescence signal for detection.

Strong fluorescence signals also endow CRISPR-AMPED the compatibility with an array of detection modalities (Figure 1C). For example, in addition to employing a conventional benchtop real-time PCR machine for detection, we demonstrate that a custom-engineered and smartphone-attached fluorescence imaging module can achieve robust detection. Such a compact detection modality can elevate the POC-amenability of CRISPR-AMPED. Moreover, we show that a commercial microfluidic digital chip can be employed to transform CRISPR-AMPED from a bulk-based output to a digital output, where the concentration of the protein target can be quantified by the number of fluorescent digital reaction wells in the chip. Taken together, CRISPR-AMPED offers a means for sensitive, specific, rapid, quantitative, and versatile detection of protein targets.

### Testbed Model for CRISPR-AMPED

We selected IL-8 as the model target for developing and demonstrating CRISPR-AMPED. IL-8 is a well-established inflammatory biomarker with documented relevance to cancer^43-45^, chronic inflammatory diseases^46, 47^, pulmonary diseases^48^, cardiovascular disease^49, 50^, and Alzheimer’s diseases^51, 52^. In this work, we spiked IL-8 in 20 % fetal bovine serum to simulate clinically relevant sample matrices. For constructing the PEA probe in this work, we repurposed the oligo sequences used in our previous CRISPR/Cas-based nucleic acid detection assays^34^. Based on which, we directly adopted the RPA primers and crRNA for amplifying the resulting DNA template (Table S1). We also note that our crRNA was designed without the need to target the protospacer adjacent motif (PAM), as we had empirically determined previously that such a PAM-free crRNA led to more sensitive single-step RPA-CRISPR/Cas12a reaction. We began with a baseline condition of magnetic proximity extension – 5 million beads per reaction, 60 min of IL-8 capturing, 60 min of binding with 500 pM PEA probe, and 10 min of extension with 0.3 U/µL *Bst* polymerase – and a baseline condition of RPA-CRISPR/Cas12a reaction – 160 nM RPA primers and 640 nM Cas12a-crRNA complex at 37 °C for 60 min. At this baseline condition, CRISPR-AMPED could detect 100 pg/mL and 1 pg/mL of IL-8 with strong normalized end-point fluorescence signals (see Methods for calculation; hereafter as “fluorescence signals”) and pronounced fluorescence amplification curves. Moreover, for the no-target control, CRISPR-AMPED output only a weak fluorescence signal and a flat fluorescence curve (Figure S2A). These results demonstrate the feasibility and establish the baseline condition for CRISPR-AMPED for detecting IL-8.

### Optimization of CRISPR-AMPED

We then systematically adjusted several parameters for magnetic proximity extension in an effort to accelerate CRISPR-AMPED while retaining strong fluorescence signals for IL-8 detection. To this end, we first found that we could shorten the time for capturing IL-8 from 60 min to 15 min and still retain robust fluorescence signal from both 100 pg/mL and 1 pg/mL of IL-8 (Figure 2A, S2B, and S3A). Then, using 15 min IL-8 capturing time, we saw decreased fluorescence signals from IL-8 when we shortened PEA probe binding time, presumably due to decrease amounts of PEA probes bound to the target protein (Figure S2C and S3B). We therefore still proceeded with 60 min as PEA probe binding time. We next increased the PEA probe concentration and found that doubling the PEA probe concentration from 500 pM to 1 nM enhanced the fluorescence signal from 1 pg/mL IL-8 but further increasing the PEA probe to 2 nM led to unwanted fluorescence signals from the NTC (Figure 2B, S2D, and S3C). We finally explored different concentrations for *Bst* polymerase and different extension times and found that 0.3 U/µL *Bst* polymerase achieved the strongest fluorescence signals from 100 pg/mL and 1 pg/mL of IL-8 and the weakest background fluorescence from the NTCs (Figure S2E and S3D). However, the extension time required careful tuning with longer extension times (20 min) increased background noise, whereases shorter times (5 min) reduced the fluorescence signal. A 10 min extension time provided an optimal balance, maintaining high fluorescent signal intensity with minimal background signals (Figure 2C, S2F, and S3E). Based on these results, we showed that we could speed CRISPR-AMPED by 45 min by shortening the time for capturing IL-8. More broadly, these results also underscore the relevant parameters for further optimizing CRISPR-AMPED.

**Figure 2:**
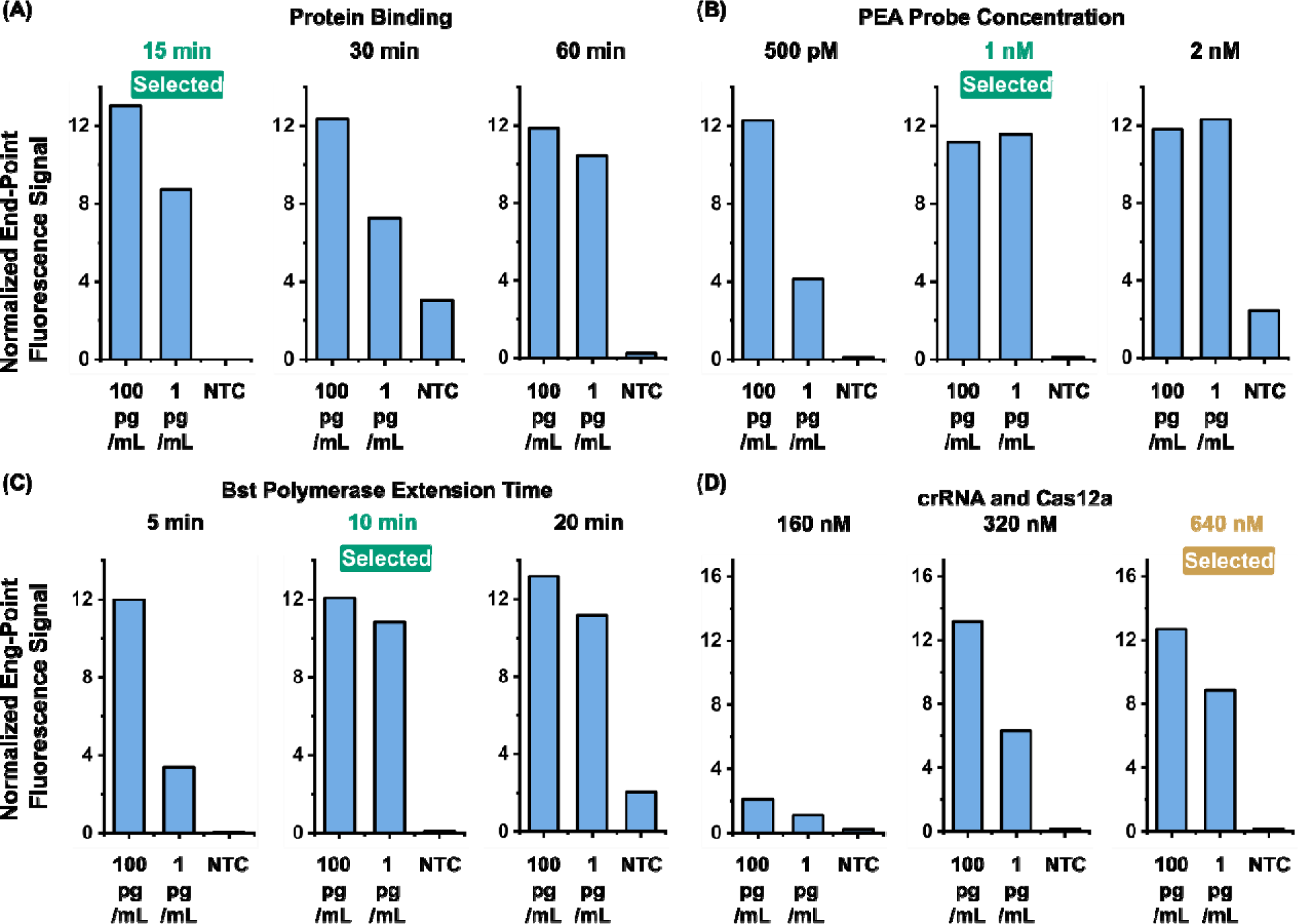
Key parameters for optimizing CRISPR-AMPED. (A) Reduction of protein binding time from 60 min to 15 min maintained high fluorescence signals for both 100 pg/mL and 1 pg/mL IL-8 targets while keeping in background signals from the NTC low. (B) Optimization of PEA probe concentration identified 1 nM as the optimal concentration. Lower concentrations (500 pM) reduced fluorescence signals for 1 pg/mL IL-8, while higher concentrations (2 nM) caused unwanted background signals. (C) Selection of the *Bst* polymerase extension time at 10 min balanced fluorescence signal and background noise. A shorter extension time (5 min) resulted in reduced signals from 1 pg/mL IL-8, while a longer extension time (20 min) led to increased background signals. (D) Tuning of crRNA and Cas12a concentrations in the RPA-CRISPR/Cas reaction to 640 nM ensured sufficient Cas12a activation, generating the highest fluorescence signals for both 100 pg/mL and 1 pg/mL IL-8 targets while minimizing background signals from the NTC. Error bar depicts standard deviations.

We subsequently investigated key parameters of our RPA-CRISPR/Cas12a reaction to ensure sensitive detection of IL-8. Specifically, when we increased the reaction temperature from 37 °C to 39 °C and 42 °C, we observed an unwanted increase in the fluorescence signals from the NTCs (Figure S4A and S5A), prompting us to select 37 °C as the reaction temperature. We next assessed three RPA primer concentrations at 80 nM, 160 nM, and 320 nM, and saw that while these three concentrations produced comparable fluorescence signals, 160 nM yielded the strongest fluorescence signals from both IL-8 samples and the lowest fluorescence signal from the NTC (Figure S4B and S5B). We finally decreased the crRNA-Cas12a concentrations (at a fixed 1:1 ratio between crRNA and Cas12a) from 640 nM to 320 nM and 160 nM. We found that the two lower crRNA-Cas12a concentrations weakened the fluorescence signals from 1 pg/mL IL-8 (Figure 2D, S4C, and S5C), likely due to insufficient activated Cas12a. In summary, we adopted 37 °C, 160 nM RPA primers, and 640 nM crRNA-Cas12a as the optimal condition for the RPA-CRISPR/Cas12a reaction of CRISPR-AMPED.

### Performance of CRISPR-AMPED

For demonstrating the performance of CRISPR-AMPED, we first showed that it could specifically detect IL-8. Here, we tested CRISPR-AMPED against 100 pg/mL of IL-8, IL-6, and TNF-α and compared their fluorescence signals with that of the NTC, all in triplicates (Figure 3A and S6). Whereas IL-8 resulted in a robust fluorescence signal above the threshold (i.e., mean + 3 × standard deviation; red dashed line in Figure 3A) set from the background fluorescence signal of the NTC, IL-6 and TNF-α both failed to register fluorescence signals above the threshold. These results illustrate the specificity of CRISPR-AMPED for IL-8 detection.

**Figure 3.**
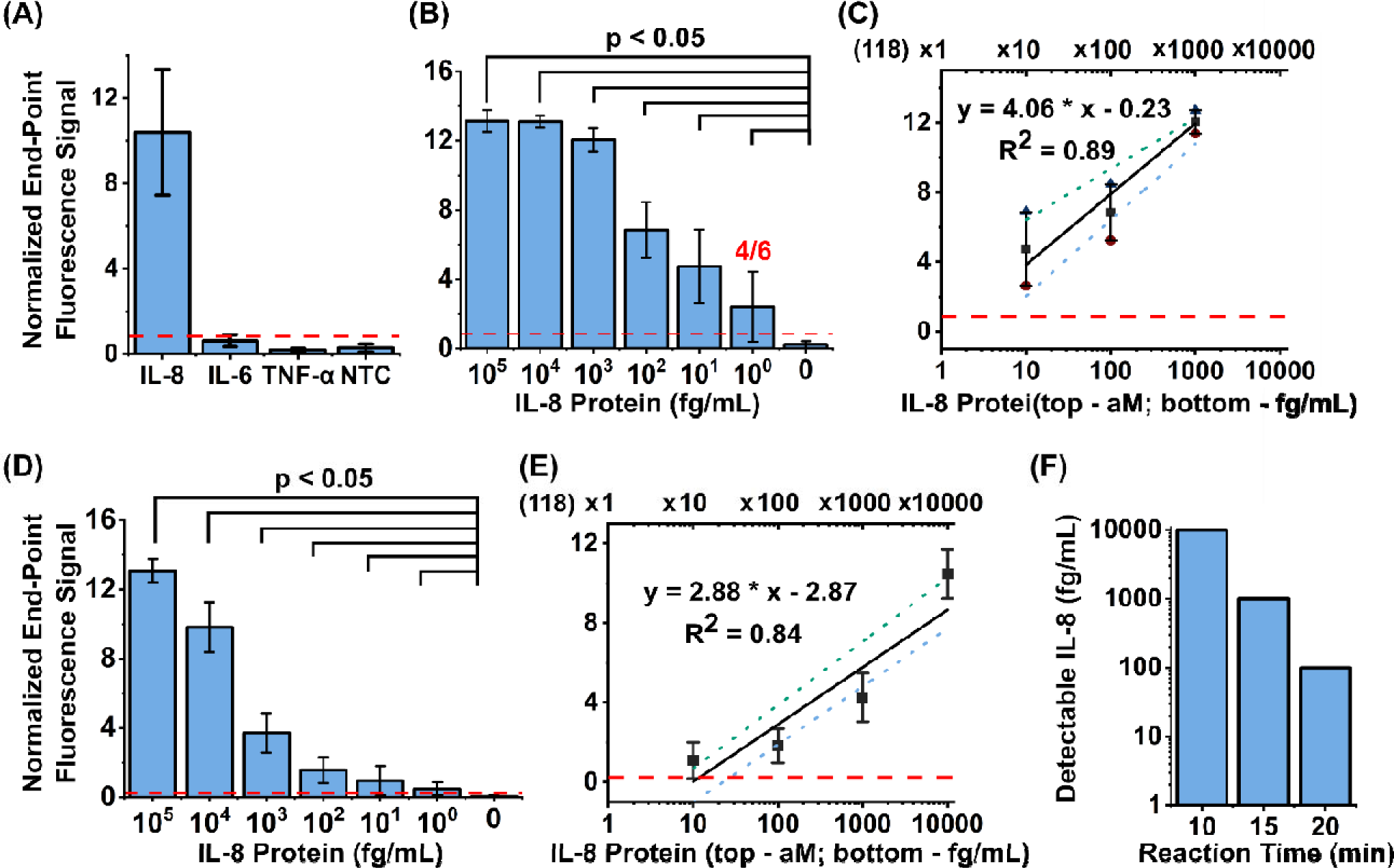
Sensitivity and specificity of CRISPR-AMPED. (A) The specificity of CRISPR-AMPED was evaluated by spiking in different proteins, IL-8, IL-6, and TNF-α at a high concentration of 100 pg/mL in 20% fetal bovine serum. (n = 3). Only IL-8 targets showed a detectable end-point signal of 10.40 above the threshold of 0.85 (red dashed line). (B) The sensitivity of the assay with 60 min reaction time was tested ranging from 1 to 10^5^ fg/mL, corresponding to 11.8 nM to 118 aM (n = 6). The assay showed the ability to detect all concentrations (p < 0.05), but the detection limit was defined as 10 fg/mL due to the 4/6 of detecting 1 fg/mL. (C) Linear regression between the IL-8 protein concentration and the end-point signal (Figure 4B) was performed for quantification in bulk assay (n = 6) for the 60 min reaction. Two more fittings were performed to incorporate the LOD range at mean ± SD (Green dotted line – mean + SD, blue dotted line – mean – SD, same for C and E). (D) The sensitivity of the assay with 30 min reaction time was tested, which showed the ability to detect all concentrations (p < 0.05), but the detection limit was defined as 10 fg/mL due to the 4/6 of detecting 1 fg/mL. (E) Similar linear regression between the IL-8 protein concentration and the end-point singal was performed for quantification in bulk assay (n = 6) for the 30 min reaction (F) Detectable IL-8 concentration was shown at different reaction times (10, 15, 20, and 30). Error bar depicts standard deviations.

We next demonstrated that CRISPR-AMPED can achieve highly sensitive detection of IL-8. To do so, we tested CRISPR-AMPED against serial dilutions of 10^5^ to 1 fg/mL IL-8, which is equivalent to 11.8 nM to 118 aM. The average fluorescence signals (n = 6) across all tested concentrations of IL-8 were detectable above the NTC threshold (red dashed line in Figures 3B and S7). Using Students t-tests as the metric for evaluating the sensitivity, we found that all tested concentrations of IL-8 produced significantly greater fluorescence signals than those of the NTC (p < 0.05). Nevertheless, we note that 2 out of the 6 replicates of 1 fg/mL IL-8 failed to produce detectable fluorescence signals. We further calculated the limit of detection (LOD, in molar concentration) via linear regression analysis. Here, in addition to fitting the mean fluorescence signals, we also performed fitting for mean – 1 × standard deviation and mean + 1 × standard deviation as the upper and lower bands (Figure 3C). Based on linear regression analysis, we calculated our LOD as 217.9 aM, and upper and lower bands as 637.3 aM and 4.6 aM, respectively. These results strongly support that CRISPR-AMPED can achieve aM-level detection of IL-8. Since we measured the fluorescence signals from the RPA-CRISPR/Cas12a reaction in real-time, we further investigated the sensitivity of CRISPR-AMPED as a function of the reaction time. We found that RPA-CRISPR/Cas12a reaction time could be reduced to 30 min while retaining the capability of detecting 10 fg/mL IL-8 and 4 out of the 6 replicates of 1 fg/mL IL-8 (Figure 3D and S8). Based on the linear regression analysis with 30 min reaction, we calculated the LOD and the upper and lower bands as 1.4 pM, 3.2 pM, and 0.9 pM, respectively (Figure 3E). The results demonstrated that 30 min RPA-CRISPR/Cas12a reaction time maintained the sensitivity of CRISPR-AMPED within single-digit pM range. Further reducing the reaction time could result in a trade-off with the lowest experimentally detectable concentration. That is, when we reduced the reaction time to 20, 15, and 10 min, we found that the lowest experimentally detectable concentration further raised to 10^2^, 10^3^, and 10^4^ fg/mL, respectively (Figure 3F). More broadly, these results illustrate that while the assay time for CRISPR-AMPED could be reduced, the trade-off with assay sensitivity must be considered.

### Portable Detection via a Smartphone-Attached Fluorescence Imaging Module

POC diagnostics are increasingly important in clinical and field settings, where rapid, accurate, and accessible detection of biomarkers is essential for timely medical decision-making^53-55^. Traditional laboratory-based fluorescence detectors, while highly sensitive, are often bulky, expensive, and require specialized infrastructure and trained personnel, which limits their use in decentralized and resource-limited environments. To address these limitations and enhance the practicality of CRISPR-AMPED for POC applications, we developed a portable smartphone-attached fluorescence imaging module capable of detecting the strong fluorescence signals generated by the CRISPR-AMPED assay. For this demonstration, we custom engineered the fluorescence imaging module by 3D printing a compact housing with a holder for reaction tubes and assembling within this housing a LED, a fluorescence filter cube, and control circuitry (Figure 4A). We designed the housing such that it could be directly attached to a smartphone while holding the reaction tubes in focus and within the view field of the smartphone camera. Of note, this smartphone-attached fluorescence imaging module supported only end-point detection, as it lacked built-in heating capability. Nevertheless, the operation was simple. Upon the completion of RPA-CRISPR/Cas12a reaction, up to three reaction tubes could be directly mounted in the tube holder before fluorescence images could be directly acquired using the built-in camera app of the smartphone.

**Figure 4.**
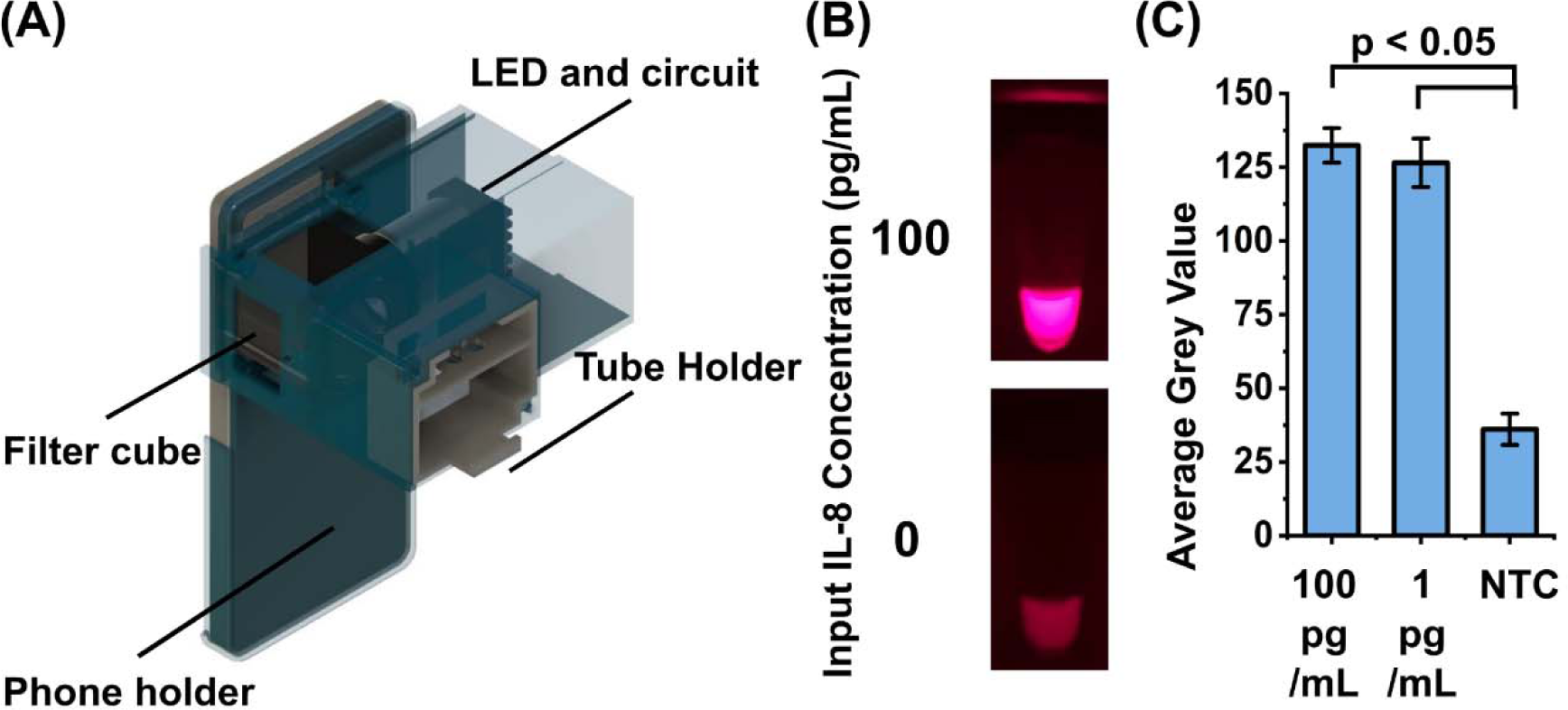
Smartphone-based detection device. (A) The device was 3D printed to enclose the optical module (red light-emitting diode (LED) and circuit, and filter cubes) and enable the mounting of the smartphone and the sample tubes. (B) With the red LED as the excitation resource and the excitation filter to illuminate the reaction tubes, two representative tube images were taken using the smartphone with emission filter, shown with 100 pg/mL and 0 pg/mL IL-8 concentration. (C) The smartphone detection showed distinguishable signal from 100 pg/mL and 1 pg/mL IL-8 target to NTC (n = 3, p < 0.05). Data represented in (C) as mean ± SD.

Even when coupled to portable detection via the smartphone-attached fluorescence imaging module, CRISPR-AMPED could still achieve robust detection. For illustration, we tested this portable detection version of CRISPR-AMPED against 100 pg/mL IL-8, 1 pg/mL IL-8, and NTC. We observed strong fluorescence from both 100 pg/mL and 1 pg/mL IL-8 and noticeably weaker fluorescence from the NTC (Figure 4B). Statistical analysis supported that 100 pg/mL and 1 pg/mL IL-8 indeed yielded significantly greater fluorescence intensities than the NTC (Figure 4C; n = 3; p < 0.05). These results illustrate that CRISPR-AMPED could be executed without bulky fluorescence detectors, thereby clearing a notable barrier for potential point-of-care use.

### Precise and Quantitative Detection via Microfluidic-based Digitization

Digitization of CRISPR/Cas-based assays represents an emerging detection approach with the potential to improve assay sensitivity, dynamic range, precision, and speed^38, 42, 56, 57^. We were thus motivated to investigate the feasibility and potential of digital CRISPR-AMPED. To do so, we adopted the RPA-CRISPR/Cas12a portion of CRISPR-AMPED for digitization. Specifically, upon magnetic proximity extension, we aliquoted 10% of magnetic beads (along with the extension product on their surfaces), mixed them with RPA-CRISPR/Cas12a reaction, and loaded the mixture into a commercial microfluidic digital chip with 20,000 0.7-nL reaction wells (i.e., QuantStudio 3D Digital PCR 20K Chip). Building upon our previous digital CRISPR/Cas assays^38, 56^, we incorporated Tween-20 and BSA as additives to the reaction mixture to facilitate its digitization into individual reaction wells. After 30-min incubation, we employed fluorescence microscopy to detect fluorescence from the reaction wells. The reaction wells with strong fluorescence – the positive reaction wells – were correlated with the presence of IL-8.

We tested the performance of digital CRISPR-AMPED by detecting IL-8 from 10^5^ to 10^0^ fg/mL. As we decreased the IL-8 concentration, we indeed detected fewer positive reaction wells, providing a strong indicator for successful digitization of CRISPR-AMPED (Figure 5A). Importantly, across the 5-log IL-8 concentration range we tested, we observed a linear correlation between the IL-8 concentration and the number of positive reaction wells, as well as small standard deviations at all IL-8 concentrations (Figure 5B). These results demonstrated that magnetic beads and the magnetically bound DNA extension products in the reaction mixture could be appropriately digitized. The linear correlation – unlike the correlation that we observed for bulk-based CRISPR-AMPED, which plateaued at high IL-8 concentrations – was particularly suitable for linear regression analysis. Based on linear regression analysis, we found that the LOD was 309.9 aM. Moreover, because of the small standard deviations, the upper and lower bands were 338.5 aM and 308.8 aM. These results revealed that digital CRISPR-AMPED could offer more precise quantitation than its bulk-based counterpart. However, we note that we currently observed only 3 orders of magnitude change in the number of positive reaction wells from 4 orders of magnitude change in the IL-8 concentration, suggesting that we currently could not detect all individual IL-8 molecules. Nonetheless, these results still demonstrated that digital CRISPR-AMPED afforded a wider dynamic range than bulk-based CRISPR-AMPED with 60-min RPA-CRISPR/Cas12a reaction time and achieved a better sensitivity than bulk-based CRISPR-AMPED with 30-min RPA-CRISPR/Cas12a reaction time.

**Figure 5:**
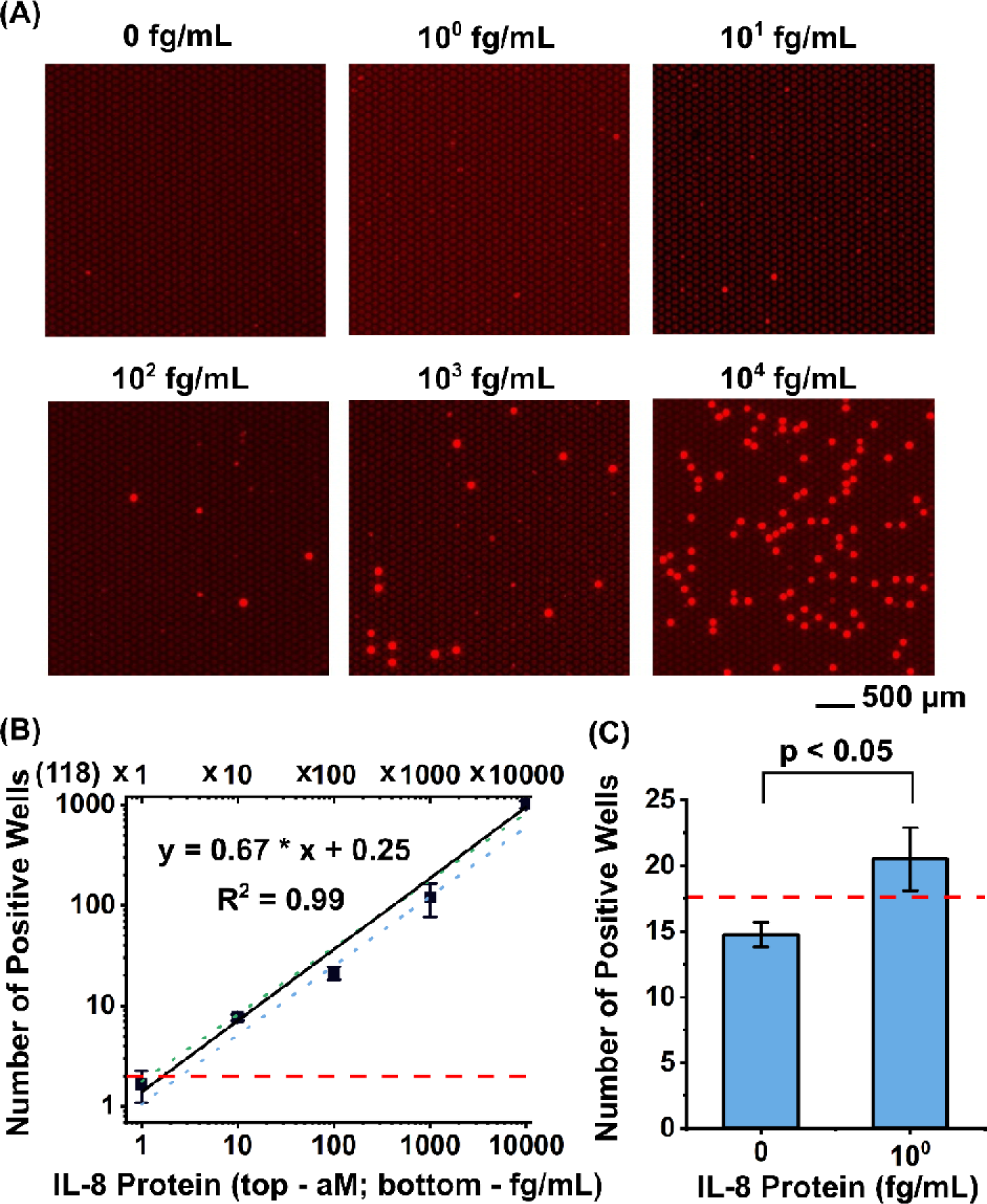
CRISPR-AMPED in QuantStudio Digital Chip. (A) Upon digitizing IL-8 target concentrations ranging from 10^0^ to 10^5^ fg/mL, strong fluorescence signal in each digital well was observed with only a few non-specific wells in NTC, shown with representative images. (B) Linear regression between the IL-8 protein concentration and number of positive wells was performed for quantification in digital chip (n = 3) after 30 min reaction. In the digital format, the LOD was 309.9 aM and ranged between 308.8 – 338.5 aM. (C) 10^0^ fg/mL was significantly different from NTC (p < 0.05) when using 100% of the beads captured with protein target (n = 3). Data represented as mean ± SD. Red dashed line represents signal threshold at mean + 3SD of the NTCs.

In digital CRISPR-AMPED, the number of magnetic beads analyzed in microfluidic digital chips could be adjusted to alter the detection performance. This strategy is consistent with digital ELISA. As an example, we experimented with using 100% of the magnetic beads as a means to boost the detection sensitivity. Specifically, when we used 100% of the magnetic beads to detect 1 fg/mL IL-8, we were able to observe statistically significant increase in the number of positive reaction wells when compared to the NTC (Figure 5C; n = 3; p < 0.05). On the other hand, when we used 100% of the magnetic beads, we observed that the number of positive reaction wells started to plateau at or near the maximum number of reaction wells, thus narrowing the dynamic range. These results illustrate the relationships between the number of magnetic beads, the sensitivity, and the dynamic range in digital CRISPR-AMPED.

## Discussion

By combining magnetic proximity extension and RPA-CRISPR/Cas12a reaction, CRISPR-AMPED achieved attomolar level detection of IL-8, placing it among the best CRISPR/Cas-based protein detection^24-27^. Furthermore, CRISPR-AMPED demonstrated greater sensitivity than several other detection methods. Indeed, the LOD demonstrated in this work was > 6000-fold lower than that of conventional ELISA^58^ and > 60-fold lower than the LOD of solid-phase PLA^58^ and PEA^28^. In fact, CRISPR-AMPED was also 6-fold more sensitive than our previously reported MagPEA with PCR-based DNA amplification and detection^33^ (Table 1). The superior sensitivity may be due to the dual advantages of reduced background noise due to the use of magnetic beads in the PEA process and the enhanced sensitivity conferred by the RPA-CRISPR/Cas12a reaction. Finally, the LOD of CRISPR-AMPED was only two-fold higher than that of digital ELISA^59^ (Table 1). Nonetheless, CRISPR-AMPED offers a significant advantage in versatility by utilizing nucleic acid barcodes instead of colorimetric beads, which could broaden its applicability in diverse diagnostic settings.

**Table 1.** LOD of detecting IL-8 proteins among different assays.

| Different assays | LOD (aM) |
| --- | --- |
| ELISA | $1.99 \times 10^6$<sup>58</sup> |
| Solid-phase PLA | $1.99 \times 10^4$<sup>58</sup> |
| PEA | $2.36 \times 10^4$<sup>28</sup> |
| MagPEA | $1.88 \times 10^3$<sup>33</sup> |
| Digital ELSIA | 177.1<sup>59</sup> |
| CRISPR-AMPED (bulk) | 217.9 |
| CRISPR-AMPED (Digital) | 309.9 |

Through the development of CRISPR-AMPED, we demonstrated the synergy between magnetic proximity extension and RPA-CRISPR/Cas12a reaction, suggesting that RPA-CRISPR/Cas12a reaction could supplant the role of PCR. Indeed, RPA-CRISPR/Cas12a reaction obviates thermocycling and provides an edge in assay speed, especially when performed using benchtop instrumentation. More broadly, our work points to the potential of exploring other isothermal NAATs such as RPA^37, 60^ (without CRISPR/Cas) and LAMP (loop-mediated isothermal amplification)^61, 62^ as a companion for magnetic proximity extension assay.

Because our work represents the first time that magnetic proximity extension and RPA-CRISPR/Cas12a reaction was combined, we recognize and point out many directions for further advances. Future work should focus on expanding the range of detectable protein targets and optimizing the assay for broader clinical applications^63-69^, as well as the capability of detecting protein targets in different sample types (i.e., saliva, blood, and other bodily fluids)^70^. Further assay optimization – such as developing a single-step reaction that encompasses probe extension, RPA, and CRISPR/Cas reaction – also warrants investigation. For the bulk format, expanding the current smartphone-based fluorescence imaging module to include temperature control and integrating a heater could improve POC performance. Alternatively, adaption to magnetofluidics technology, which has been employed to demonstrate CRISPR/Cas-based nucleic acid detection^71^ and immunoPCR-based protein detection^72^ could also deliver a fully integrated device that is amenable for point-of-care testing^73, 74^. For the digital format, digitization can also open the door to multiplexing^56^. Implementation of digital CRISPR-AMPED in custom microfluidic chips housing more reaction wells with smaller volumes may widen the dynamic range while shortening the reaction time. Moreover, implementation of digital CRISPR-AMPED in our recently reported smartphone-based portable digital device^75^ can make digital CRISPR-AMPED more amenable for POC use. Alternatively, it is possible to implement digital CRISPR-AMPED within microvalve-based programmable droplet microfluidics technology^76, 77^ as a means to improve the dynamic range and scalability.

## Conclusion

In this work, we have successfully developed and validated CRISPR-AMPED, a novel immunoassay system combining magnetic PEA with RPA-CRISPR/Cas12a-assisted detection for highly specific and sensitive protein detection. CRISPR-AMPED integrates the advantages of magnetic beads in PEA process to reduce the background signal and the boosted sensitivity from the combination of RPA and CRISPR/Cas12a. Moreover, CRISPR-AMPED was optimized with shorter assay time and high sensitivity as demonstrated with the IL-8 protein biomarker in fetal bovine serum, making it well-suited for rapid and efficient diagnostics. Specificity was tested by spiking high concentrations of IL-6 and TNF-α, affirming the assay’s robustness against non-specific binding. CRISPR-AMPED achieved a limit of detection of 217.9 aM, demonstrating over 1,000-fold^24-27^ greater sensitivity than other antibody-assisted CRISPR/Cas-based strategies and demonstrated higher sensitivity compared to standard methods such as ELISA^58^, PLA^58^, PEA^28^, and MagPEA^33^, and comparable performance to digital ELISA^59^. The development of a smartphone-based detection device further enhances its potential for POC applications, providing a user-friendly and portable solution suitable for diverse diagnostic settings. Moreover, in digital format, CRISPR-AMPED extends dynamic range and enhances quantitation precision. Overall, the CRISPR-AMPED immunoassay represents a significant advancement in protein detection technology, combining rapid, accurate, and highly sensitive detection with versatile applicability. These advancements position CRISPR-AMPED as a transformative tool for protein biomarker detection, with promising implications for biomedical research, disease diagnostics, and future POC testing.

## Material and Methods

### Materials

The 1-μm (diameter) magnetic beads (Dynabeads MyOne Carboxylic Acid), 40kDa Zeba columns (0.5 mL/75 μL), and 7kDa Zeba columns were purchased from Thermofisher Scientific (Waltham, MA) and stored at -4 °C. The sulfo-SMCC and fetal bovine serum were purchased from Thermofisher Scientific (Waltham, MA) and stored at -20 °C. 1-Ethyl-3-(3-dimethylaminopropyl) carbodiimide (EDC) and N-hydroxy succinimide (NHS) used for magnetic beads-antibody coupling were purchased from Millipore Sigma (St. Louis, MO) and stored at -20 °C. Recombinant human IL-8 protein and human IL-8 polyclonal antibody were ordered from R&D systems (Minneapolis, MD) and stored at -20 °C. Bst 2.0 warmstart DNA polymerase was purchased from New England Biolabs (Ipswich, MA). A comprehensive inventory of reagents required for the preparation of various buffers used in the magPEA assay can be found in our previously published paper^33^. The TwistAmp Basic and TwistAmp exo RPA assay kits were purchased from TwistDx Limited (Maidenhead, UK) and stored at -20 °C. All Cas12a-guide RNA (crRNA) and DNA oligonucleotides including PEA probes, primers, ssDNA fluorescence reporters were purchased from integrated DNA Technologies (IDT; Coralville, IA) and stored at -20 °C. The crRNA sequence was modified with IDT’s proprietary AltR1 and 3’ AltR2 modifications. ssDNA fluorescence reporters were labeled with Alexa 647 and IAbRQSp. Lyophilized crRNAs, ssDNA reporters, and primers were reconstituted in nuclease-free water (Quality Biological Inc, Gaithersburg, MD) at 20 µM, 100 µM, 10 µM, respectively. EnGen Lba (Lachnospiraceae bacterium ND 2006) Cas12a (Cpf1) (100 µM) was purchased from New England BioLabs (Ipswich, MA) and stored at -20 °C. Bbovine serum albumin (BSA; 20 mg/mL) were purchased from New England BioLabs (Ipswich, MA) and stored at -20 °C. Tween 20 was purchased from MilliporeSigma (St. Louis, MO) and used without further purification.

### Antibody Conjugation onto Magnetic Beads

EDC/NHS chemistry was used to conjugate IL-8 antibodies onto magnetic beads. Briefly, 20 μL of beads (10 mg/mL) were washed in 120 μL activation buffer (100 mM MES, 0.01% Tween-20, pH 4.5), followed by resuspension in 180 μL of activation buffer. For surface activation, 20 μL of a freshly prepared solution containing 80 mg/mL EDC and 80 mg/mL NHS was added to the beads, and the mixture was allowed to react at room temperature for 15 min. After activation, the magnetic beads were washed with 120 μL activation buffer to remove any residual EDC/NHS, followed by resuspension in 15 μL activation buffer. Next, 5 μL of 1 mg/mL IL-8 antibody solution was added into the bead suspension and incubated at 4 °C overnight on an end-over-end rotator. To halt the beads coupling process, the remaining antibody supernatant was removed, and the beads were resuspended in 40 μL quenching buffer (100 mM Boric Acid, 30 mM 2-(2-aminoethoxy) ethanol, 0.01% Tween-20, pH 8.5) for 30 min. The beads were then washed with 120 μL washing buffer (10 mM Phosphate buffer, 0.05% Tween-20). Finally, beads were stored in 200 μL of storage buffer (10 mM Phosphate buffer, 0.1% BSA, 0.02% Tween-20, 0.05% sodium azide, pH 7.4).

### Antibody-Oligo Conjugation

The conjugation of thiol-modified oligonucleotides to antibody was performed as follows: 14 μL of 1 mg/mL purified antibody solution was activated by mixing with 2 μL of 3.33 mM sulfo-SMCC and incubated at 4 °C for 2 h. Meanwhile, 2.6-μL oligonucleotides at a concentration of 500 μM was reduced by adding 4.4 μL of 40 mM DTT, followed by incubating at 95 °C for 2 min and 37 °C for 1 h. Next, the activated antibody solution and reduced oligonucleotides were purified using 40kDa and 7 kDa Zeba columns respectively according to the manufacture’s instruction. After determining the concentrations on Nanodrop, the antibody was mixed with the oligonucleotides at 10×/5×/2× molar excess of oligonucleotides to antibody and incubated overnight at 4 °C for conjugation. Next, the antibody-oligo conjugates were purified using 40kDa Zeba columns to remove residual oligonucleotides. Before use, concentrations of conjugated antibody-oligo were determined using BCA assay.

### Magnetic PEA

Magnetic PEA began with blocking the antibody-coupled magnetic beads (1 mg/mL) blocked in 10% goat serum at room temperature for 1 h to minimize nonspecific adsorption. The beads were then washed twice in washing buffer (1× PBS, 0.05% Tween-20, 150 mM NaCl) before resuspending in 0.1% BSA in PBST buffer at a concentration of 0.5 mg/mL. Subsequently, the beads were aliquoted into 100 μL of serially-diluted IL-8 protein solution spiked in 20% fetal bovine serum and incubated on an end-over-end rotator for 15 min. After incubation, the beads were washed for three times, resuspended in 20-μL PEA probes in PEA buffer (1% BSA, 0.2 mg/mL ssDNA, 0.1% Tween, 150 mM NaCl, 0.05% dextran sulfate, and 5 mM EDTA) and incubated for another 1 h to form the magnetic beads-protein-PEA probe complex. The beads were then washed twice to remove the free PEA probe after immunobinding. To perform the oligo extension, the beads were first resuspended in 10 μL of extension buffer (containing 1x isothermal amplification buffer, 6 mM MgSO_4_, 1.4 mM dNTP mix, 1 unit of Bst 2.0 DNA polymerase) and incubated at 50 °C for 10 min. The extension buffer was then removed, and the beads were resuspended in 10 μL 1× PBS for downstream RPA-CRISPR/Cas12a-assisted assay.

### Assembly of the Bulk and Digital RPA-CRISPR/Cas12a-Assisted Mix

The assembly of the bulk CRISPR/Cas12a-assisted RPA mix began by resuspending each dried pellet of the TwistAmp basic reaction mix with 29.5 μL rehydration buffer to prepare a 1.7× rehydrated TwistAmp basic reaction mix. The optimized bulk RPA-CRISPR/Cas12a-Assisted mix contained the following: 1× rehydrated TwistAmp basic reaction mix, 160 nM each of RPA primers, 4 μM ssDNA fluorescence reporters, 640 nM EnGen Lba Cas12a, 640 nM crRNAs, 14 mM MgOAc, and the protein targets on the magnetic beads. All components except the protein targets on the magnetic beads and MgOAc were first assembled in 1.5 mL protein low-binding microcentrifuge tubes (MilliporeSigma, Burlington, MA) inside a PCR hood (AirClean Systems, Creedmoor, NC). After a 10 min incubation at room temperature for Cas12a-crRNA complex formation, the protein targets on the magnetic beads were mixed with MgOAc and added to the mix inside a separate biosafety cabinet (The Baker Company, Sanford, ME) to prevent any carryover contamination. When assembling the digital RPA-CRISPR/Cas12a-assisted mix for QuantStudio digital chips, we added additional 0.1% Tween 20 and 0.01 mg/mL BSA in the mix, which were added after 10 min incubation for Cas12a-crRNA complex formation. The rest of the procedures remain the same.

### Optimization of Magnetic PEA Target Preparation

In the optimization experiments for magnetic PEA target preparation, 3 protein binding time, PEA probe binding time, PEA probe concentrations, Bst DNA polymerase concentrations, and BST DNA polymerase extension time were examined. The baseline assay condition used 60 min protein binding, 60 min of PEA probe binding, 500 pM PEA probe, 0.3 U/µL Bst DNA polymerase, 10 min Bst DNA polymerase extension. The optimization of each component fixed the rest of the conditions and varied the last one. The protein binding time is tuned among 15 min, 30 min, and 60 min against 100 pg/mL and 1 pg/mL IL-8 target and NTC. The optimization of PEA probe binding time fixed the 15 min protein binding time and the rest of the conditions and varied the PEA probe binding time (15 min, 30 min, and 60 min) against 100 pg/mL and 1 pg/mL IL-8 target and NTC. The optimization of PEA probe concentration fixed the 15 min of protein binding and 60 min of PEA probe binding and the rest of the conditions and varied the PEA probe concentration (500 pM, 1 nM, and 2 nM) against 100 pg/mL and 1 pg/mL IL-8 target and NTC. The optimization of Bst DNA polymerase concentration fixed the 15 min of protein binding, 60 min of PEA probe binding, 500 pM PEA probe and the rest of the conditions and varied the Bst DNA polymerase concentration (0.06 U/µL, 0.15 U/µL, and 0.3 U/µL) against 100 pg/mL and 1 pg/mL IL-8 target and NTC. The optimization of Bst DNA polymerase extension time fixed the fixed the 15 min of protein binding, 60 min of PEA probe binding, 500 pM PEA probe and 0.3 U/µL Bst DNA polymerase and varied the Bst DNA polymerase extension time (5 min, 10 min, and 20 min) against 100 pg/mL and 1 pg/mL IL-8 target and NTC.

### Optimization of the RPA-CRISPR/Cas12a-Assisted Assay Detection

In the assay optimization experiments, 3 temperatures, and 4 concentrations of primers and crRNA&Cas12a for RPA-CRISPR/Cas12a-assisted mix were examined. The optimization of temperature fixed the 160 nM primers and 640 nM crRNA&Cas12a (Ratio 1:1) concentrations and varied the temperature (37 °C, 39 °C, and 42 °C) against 100 pg/mL and 1 pg/mL IL-8 target and NTC. The optimization of primer concentrations for RPA-CRISPR/Cas12a-assisted mix fixed the 37 °C reaction temperature and 640 nM crRNA&Cas12a (Ratio 1:1) concentrations and varied the primer concentrations (80 nM, 160 nM, 320 nM, and 640 nM) against 100 pg/mL and 1 pg/mL IL-8 target and NTC. The optimization of crRNA&Cas12a (Ratio 1:1) concentration for RPA-CRISPR/Cas12a-assisted mix fixed the 37 °C reaction temperature and 160 nM primer concentrations and varied the crRNA&Cas12a (Ratio 1:1) concentrations (80 nM, 160 nM, 320 nM, and 640 nM) against 100 pg/mL and 1 pg/mL IL-8 target and NTC.

### Smartphone-based detection device

The smartphone-based detection device was designed in Autodesk Fusion 260 (Autodesk Inc., San Rafael, CA) and 3D printed using black PLA (Prusa Research, Czech Republic). Specifically, the device is composed of an optical, a tube and a smartphone holder. Among them, the optical holder was designed to enclose the optical module and facilitate the fluorescence detection. The tube holder was designed to hold up to 3 reaction tubes and can be inserted into the optical holder to facilitate the alignment of the sample tubes. The smartphone holder was designed to mount and align the optical holder to the smartphone camera. To enable the detection of the fluorescence signal, the Cy5 fluorescence filter set (Edmund Optics, Barrington, NJ) with excitation wavelength between 604 – 644 nm and emission wavelength between 672 – 712 nm was inserted into the filter slots accompanies by a red LED (Luxeon Star LEDs, Alberta, Cananda) facing toward excitation filter. The custom LED circuit control was powered by a 9V battery. The smartphone holder was tightly mounted to the optical holder using M3 nuts and screws, which allows the smartphone camera to align well with the optical filter path. Samsung Galaxy S21 ultra (Samsung, Seoul, South Korea) was used for demonstration with the camera settings of 125 ISO, 1/15 s camera speed, 4200K WB, and 1X magnification.

### Assay Partitioning via QuantStudio Digital Chip

To perform digital magnetic PEA RPA-CRISPR/Cas12a-assisted immunoassay in QuantStudio digital chips, assembled digital RPA-CRISPR/Cas12a-assisted mix was first loaded into a commercial QuantStudio 3D Digital PCR 20K Chip v2 (ThermoFisher Scientific, Waltham, MA) by using a QuantStudio 3D Digital PCR Chip Loader and closely following the user guide. The loading process involves placing the chip, chip lid, and sample loading blade in the loader. A 15 µL Cas12a-assisted RT-RPA mix is then pipetted into the loading blade’s sample loading port. The loader moves the loading blade across the chip, dispensing the reaction mix into the wells. Immersion Fluid is added to cover the chip without touching its surface. The chip lid is pressed tightly onto the chip case, creating a sealed assembly. More Immersion Fluid is added through the fill port while allowing air to escape, until there is only a small air bubble remaining. The chip lid’s label is pressed over the fill port to establish a fully sealed chip ready for magnetic PEA RPA-CRISPR/Cas12a-assisted immunoassay in end-point format.

### End-point digital magnetic PEA RPA-CRISPR/Cas12a-assisted immunoassay

End-point digital magnetic PEA RPA-CRISPR/Cas12a-assisted immunoassay was performed by heating the chips at 37 °C using a ProFlex 2× flat PCR System (Thermo Fisher Scientific, Waltham, MA). Fluorescence signals from the chips were measured after heating using a fluorescence microscope. The chips were placed within a grid of QuantStudio 3D Digital PCR Chip Adapters on the sample block of the ProFlex system, which was set to 37 °C. QuantStudio 3D Digital PCR Thermal Pads were placed over the chip adapters, and the system lid was closed to start the reaction. The reactions were conducted for 30 min. The chips were positioned with their fill ports oriented toward the front of the ProFlex system, which was elevated to an 11° incline using the underneath QuantStudio 3D Tilt Base. After heating, each chip was transferred to a fluorescence microscope (BX51, Olympus, Japan) equipped with a collimated LED light source, an Alexa647-compatible filter cube, a 1.25× magnification objective lens, and a digital CCD camera. Fluorescence micrographs were captured from 2 distinct regions covering the whole 20,000 reaction wells, with 1 s exposure time.

### Data Processing and Statistical Analysis

For all real-time fluorescence intensity plots in the bulk assay, the fluorescence intensities at each time point (F) were normalized to the fluorescence intensity of the respective signal at t = 1 min (F_t=1_) and further subtracted 1 for zero baseline (F/F_t=1_ - 1), which can help compare different assay systems with different background intensities. For normalized end-point fluorescence signal, fluorescence intensity at 60 min (F_t=60_) was normalized to F_t=1_ and further subtracted 1 for zero baseline (F_t=60_ / F_t=1_ - 1). The fluorescence pictures obtained from the smartphone-based detection device were analyzed in ImageJ with average grey value. The fluorescence pictures of the digital assay were analyzed in ImageJ by firstly stitching the two pictures of the same chip into one and then use “adaptive threshold” to analyze the number of positive wells. All plots were plotted using OriginLab (OriginLab Corporation) and later organized with Inkscape 1.2.2 (Software Freedom Conservancy, NY, USA).

## Author Contributions

The manuscript was written through contributions of all authors. All authors have given approval to the final version of the manuscript. These authors contributed equally.

## Data Availability

All data produced in the present study are available upon reasonable request to the authors

## Acknowledgement

This research is financially supported by the National Institutes of Health (R01AI183336, R01AI181217).

## Supplementary Information

**Figure S1.**
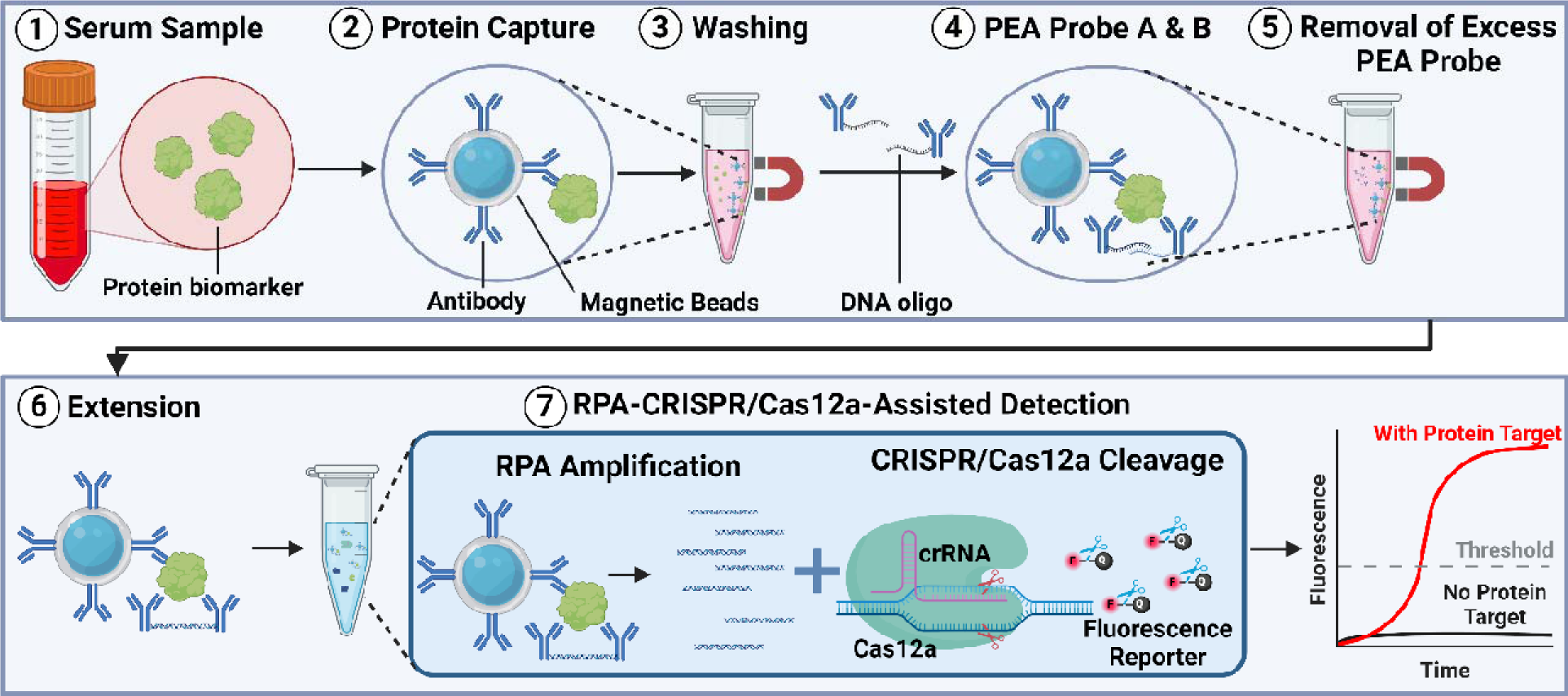
Magnetic PEA RPA-CRISPR/Cas12a-assisted immunoassay workflow. The whole process of the assay begins with the capture of protein biomarker with antibody conjugated magnetic beads (step 1 and 2). Followed with an additional washing to remove unbound proteins (step 3), the detection antibodies labeled with DNA oligos (PEA probe A & B) bind to the protein (step 4) with another washing to remove excess PEA probes (step 5). After the extension of the two DNA oligos (step 6), the DNA sequence was amplified via RPA and then detected with CRISPR/Cas12a-cleavage to yield high fluorescence signal (step 7).

**Figure S2.**
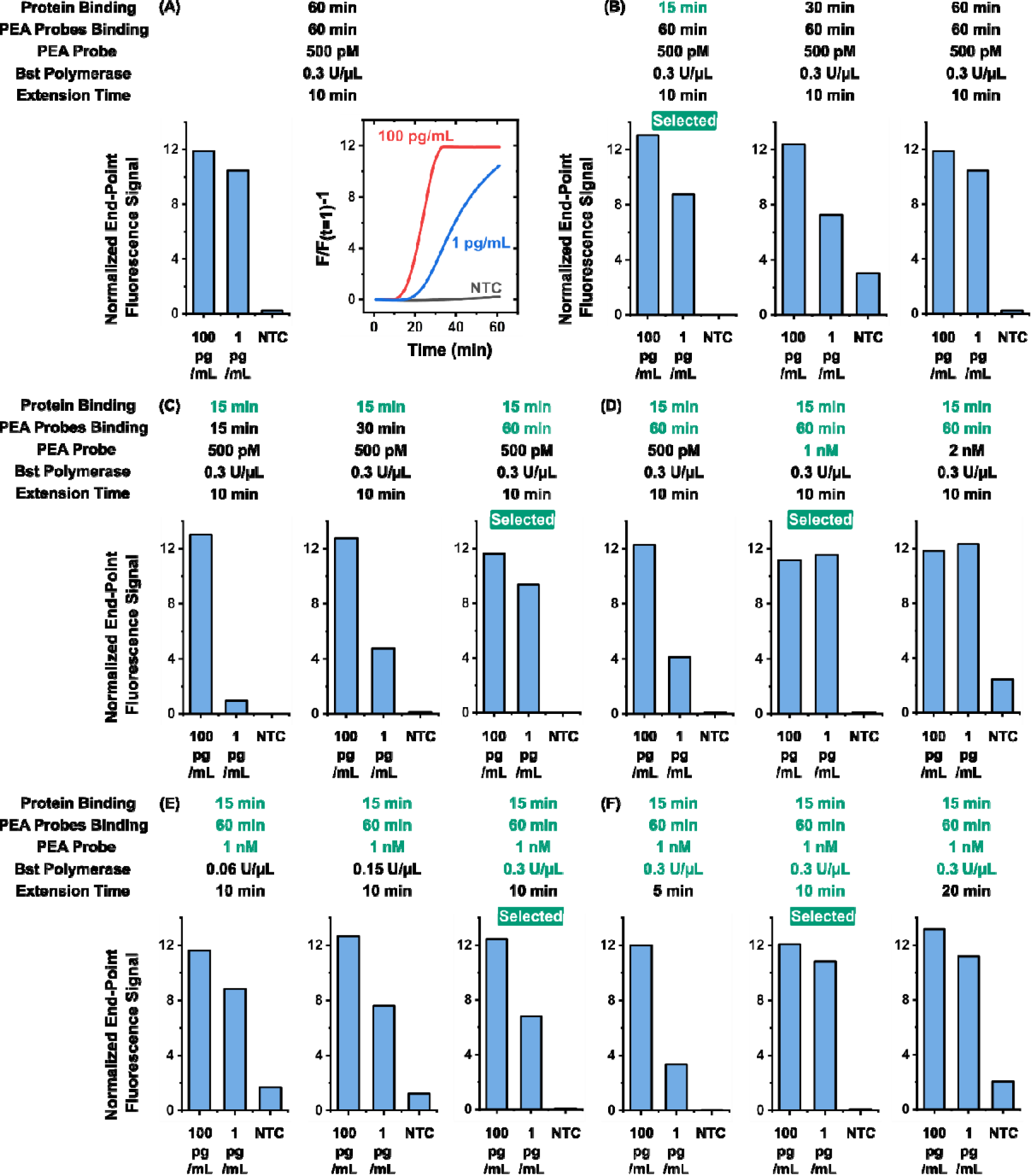
Assay optimization for magnetic PEA preparation. (A) we established a baseline assay condition with 60 min protein binding and PEA probe binding, 500 pM PEA probe, 0.3 U/µL BST, and 10 min BST extension time, and showed distinguishable signal from 100 pg/mL and 1 pg/mL IL-8 target from NTC. (B) 15 min protein binding time showed comparable end-point fluorescence signal with 30 and 60 min from 100 pg/mL (13.03) and 1 pg/mL (8.75) IL-8 target and low end-point fluorescence signal around 0 from the NTC. (C) 60 min PEA protein binding time showed the high end-point fluorescence signal from both 100 pg/mL (11.63) and 1 pg/mL (9.36) IL-8 target and low end-point fluorescence signal from the NTC (0.02). (D) 1 nM PEA probe showed the high end-point fluorescence signal from both 100 pg/mL (11.16) and 1 pg/mL (11.56) IL-8 target while maintaining a low end-point fluorescence signal from the NTC (0.13). (E) 0.3 U/µL Bst extension polymerase showed the high end-point fluorescence signal from both 100 pg/mL (12.45) and 1 pg/mL (6.81) IL-8 target while maintaining a low end-point fluorescence signal from the NTC (0.05). (F) 10 min extension time showed the high end-point fluorescence signal from both 100 pg/mL (12.08) and 1 pg/mL (10.83) IL-8 target while maintaining a low end-point fluorescence signal from the NTC (0.08).

**Figure S3.**
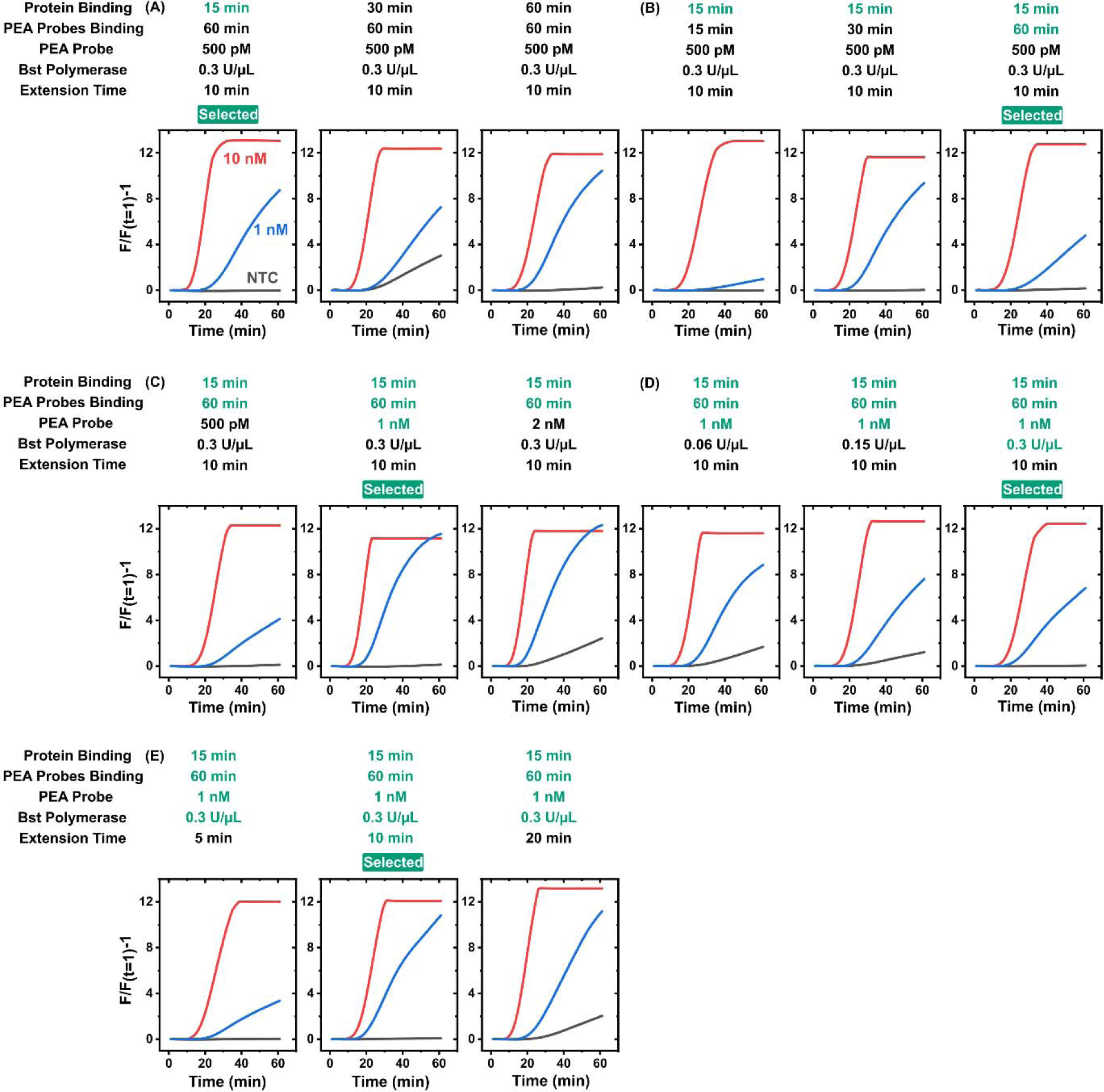
Reaction real-time curves for optimizing magnetic PEA target preparation. Here, every prepared protein target was detected by 10-µL RPA-CRISPR/Cas12a-asssited reaction mixture, with both 10 nM, 1 nM, an NTC. All assays are loaded into 0.2 mL PCR tube strips and incubated in a Bio-Rad CFX96 Touch Real-Time PCR Detection System at 37 °C for 60 min, and their fluorescence signals are measured every 1 min starting at the first min (i.e., t = 1). The fluorescence signals are extracted without baseline subtraction (i.e., under “No Baseline Subtraction” mode in the CFX Manager Software) and background subtracted normalized fluorescence signals (i.e., __) are calculated. (A) Among different protein binding time, 15 min protein binding time showed comparable fluorescence signal with 30 and 60 min from 100 pg/mL and 1 pg/mL IL-8 target and low fluorescence signal from the NTC. (B) Among different PEA probe binding time, 60 min PEA protein binding time showed the high fluorescence signal from both 100 pg/mL and 1 pg/mL IL-8 target and low fluorescence signal from the NTC. (C) 1 nM PEA probe showed the high fluorescence signal from both 100 pg/mL and 1 pg/mL IL-8 target while maintaining a low fluorescence signal from the NTC. (D) 0.3 U/µL Bst extension polymerase showed the high fluorescence signal from both 100 pg/mL and 1 pg/mL IL-8 target while maintaining a low fluorescence signal from the NTC. (E) 10 min extension time showed the high fluorescence signal from both 100 pg/mL and 1 pg/mL IL-8 target while maintaining a low fluorescence signal from the NTC.

**Figure S4.**
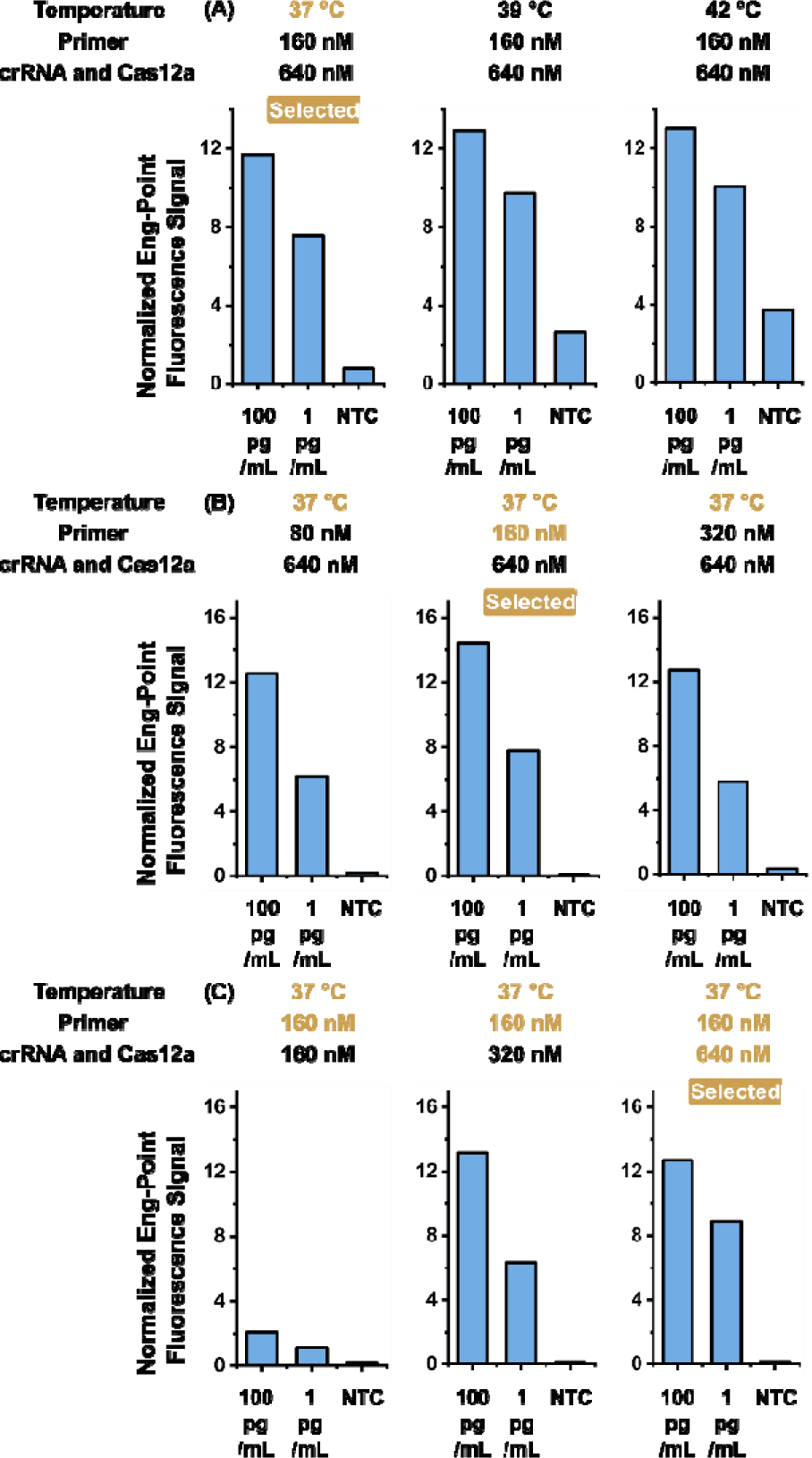
Assay optimization for RPA-CRISPR/Cas12a-assisted detection. (A) 37 °C reaction temperature results in a strong end-point signal from 100 pg/mL (11.69) and 1 pg/mL (7.58) IL-8 target, respectively, and a low end-point signal from the NTC (0.79). (B) 160 nM primer results in a strong end-point signal from 100 pg/mL (14.44) and 1 pg/mL (7.78) IL-8 target, respectively, and a low end-point signal from the NTC (0.09). (C) 640 nM crRNA-Cas12a results in a strong end-point signal from 100 pg/mL (12.70) and 1 pg/mL (8.89) IL-8 target, respectively, and a low end-point signal from the NTC (0.16).

**Figure S5.**
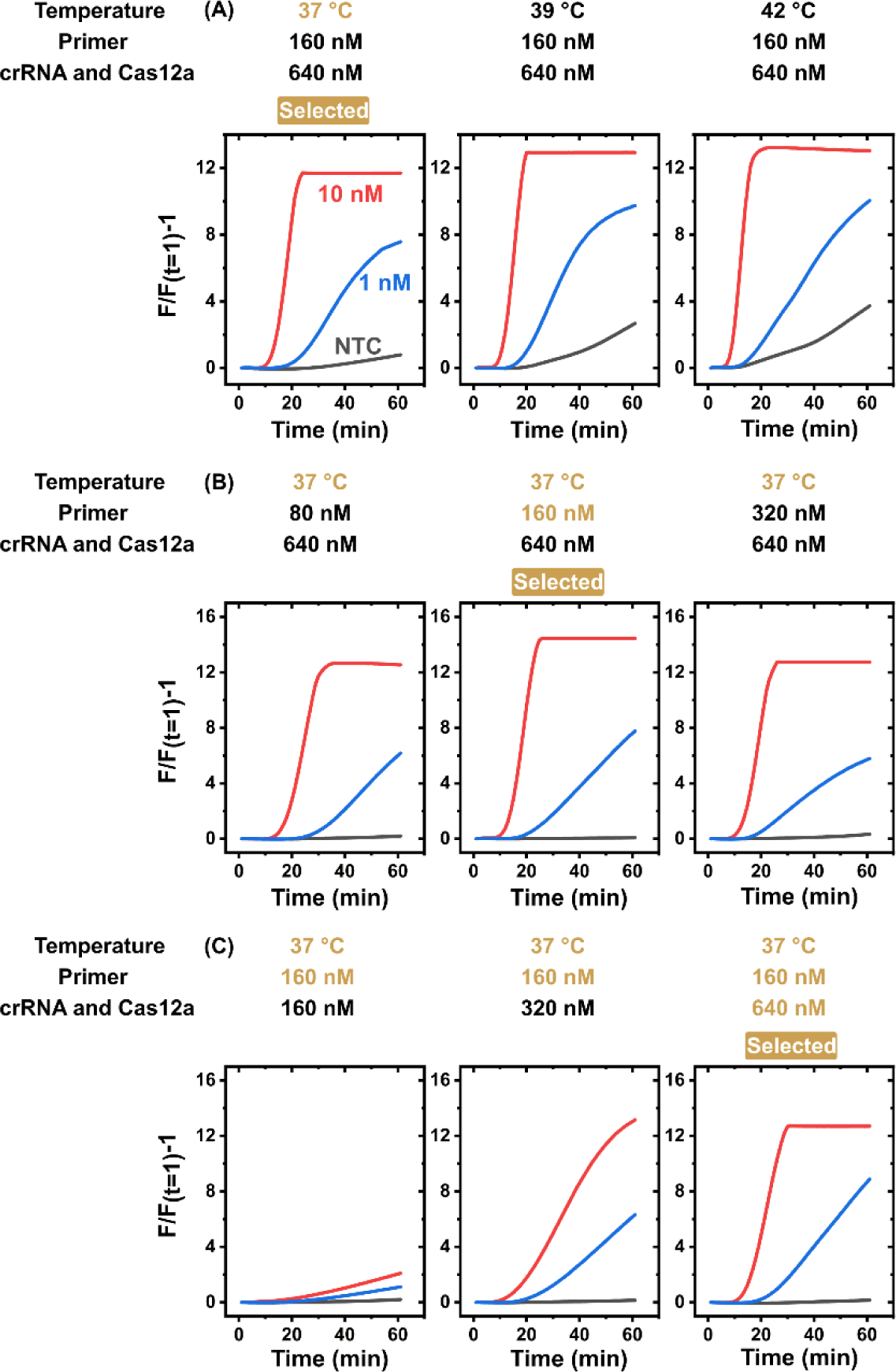
Reaction real-time curves for optimizing RPA-CRISPR/Cas12a-assisted assay. Here, every prepare protein target was prepared in the optimal condition (Figure S2) and detected by 10-µL RPA-CRISPR/Cas12a-asssited reaction mixture, with both 10 nM, 1 nM, and NTC. All assays are performed, and their background subtracted normalized fluorescence signals (i.e., ____) are calculated. As described in Figure S2. (A) 37 °C reaction temperature was chosen because it results in a strong signal from 100 pg/mL and 1 pg/mL IL-8 target, respectively, and a low signal from the NTC. (B) Among 80, 160, and 320 nM primer concentrations, 160 nM primer results in a strong signal from 100 pg/mL and 1 pg/mL IL-8 target, respectively, and a low signal from the NTC. (C) Among 160, 320, and 640 nM crRNA-Cas12a concentrations, 640 nM crRNA-Cas12a results in a strong signal from 100 pg/mL and 1 pg/mL IL-8 target, respectively, and a low signal from the NTC.

**Figure S6.**
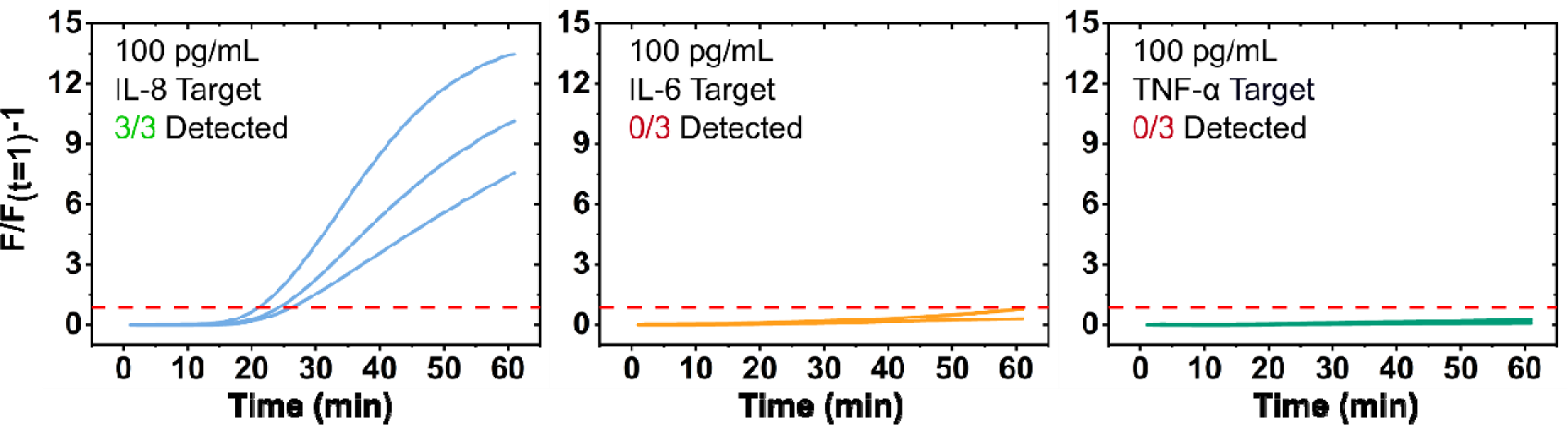
Reaction real-time curves of specificity testing of magnetic PEA RPA-CRISPR/Cas12a-assisted assay. Here, every 10-µL reaction mixture and target is prepared based on its respective optimal composition (see Figures S2 and S3) to detect different proteins, IL-8, IL-6, and TNF-α at a high concentration of 100 pg/mL in 20% fetal bovine serum. All assays are performed, and their background subtracted normalized fluorescence signals (i.e., ____) are calculated, as described in Figure S2. All concentrations are triplicated, and the number of detected replicates is recorded. Only IL-8 targets showed a detectable signal 3 out of 3 replicates above the red dash line threshold. Both IL-6 and TNF-α targets are not detectable in all 3 replicates. Red dashed line represents signal threshold at mean ± 3SD of the NTCs.

**Figure S7.**
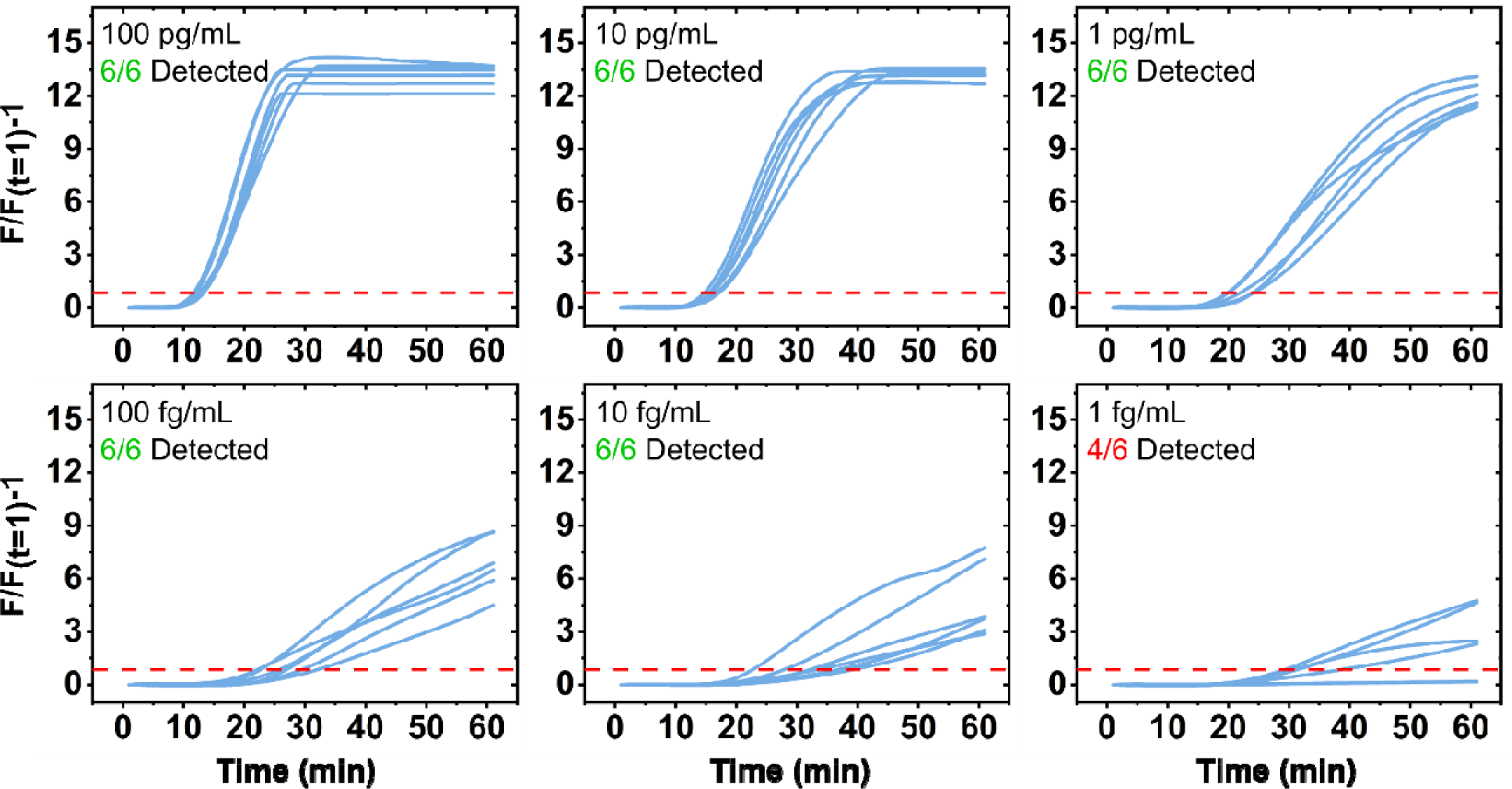
Reaction real-time curves of Magnetic PEA RPA-CRISPR/Cas12a-assisted assay for assessing assay sensitivity in 60 min reaction. Here, every 10-µL reaction mixture and target is prepared based on its respective optimal composition (see Figures S2 and S3) to detect a range of concentrations from 1 to 10^5^ fg/mL (n=6). All assays are performed, and their background subtracted normalized fluorescence signals (i.e., ___) are calculated, as described in Figure S2. The assays showed the capability to detect 10 to 10^5^ fg/mL IL-8 target in all 6 replicates above the red dash line threshold. It detects the 1 fg/mL IL-8 target in 4 out of 6 replicates. Red dashed line represents signal threshold at mean ± 3SD of the NTCs.

**Figure S8.**
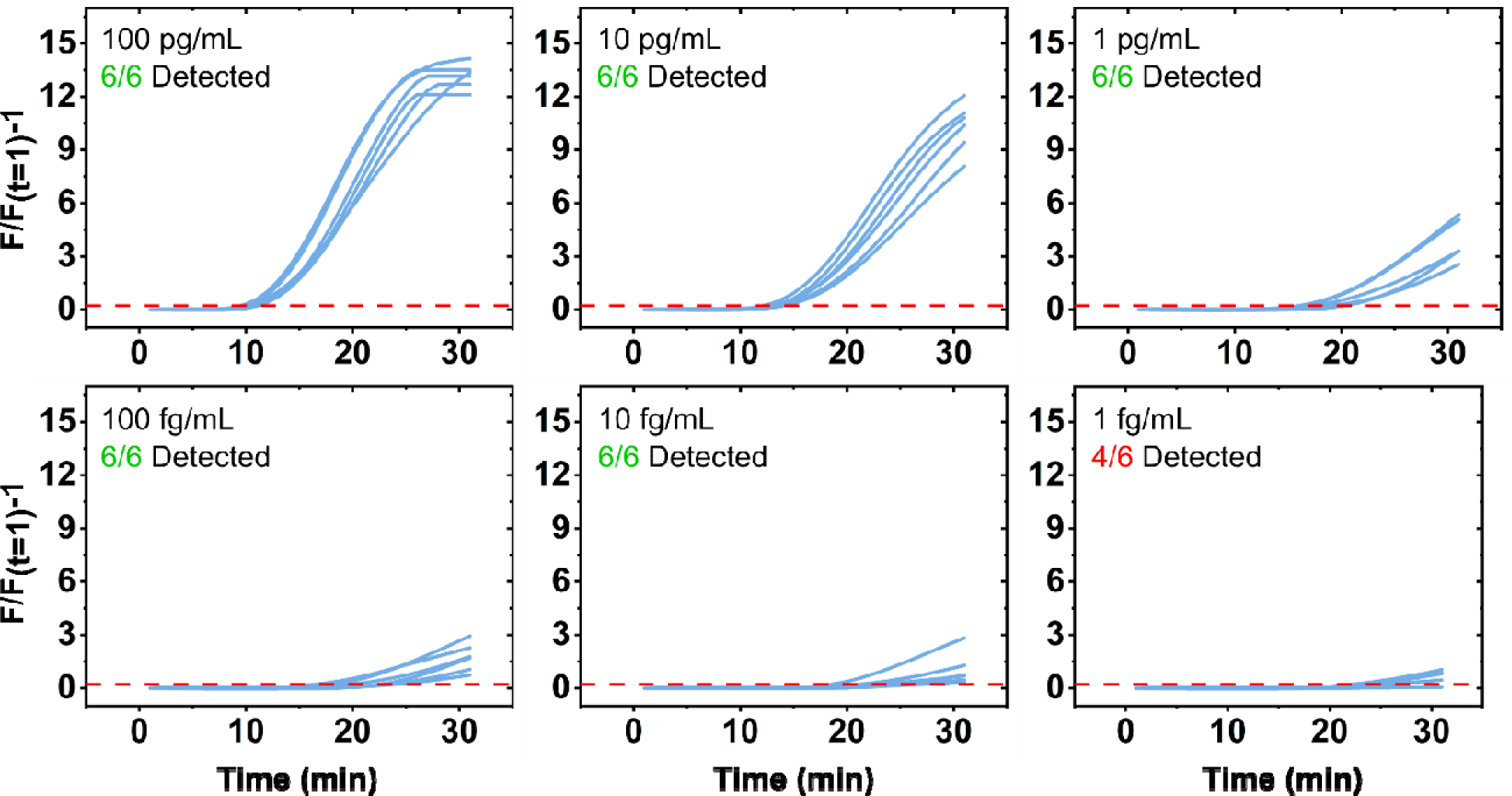
Reaction real-time curves of Magnetic PEA RPA-CRISPR/Cas12a-assisted assay for assessing assay sensitivity in 30 min reaction. Here, every 10-µL reaction mixture and target is prepared based on its respective optimal composition (see Figures S2 and S3) to detect a range of concentrations from 1 to 10^5^ fg/mL (n=6). All assays are performed, and their background subtracted normalized fluorescence signals (i.e., ___) are calculated, as described in Figure S2. The assays showed the capability to detect 10 to 10^5^ fg/mL IL-8 target in all 6 replicates above the red dash line threshold. It detects the 1 fg/mL IL-8 target in 4 out of 6 replicates, same as 60 min reaction (See Figure S5). Red dashed line represents signal threshold at mean ± 3SD of the NTCs.

**Table S1.**
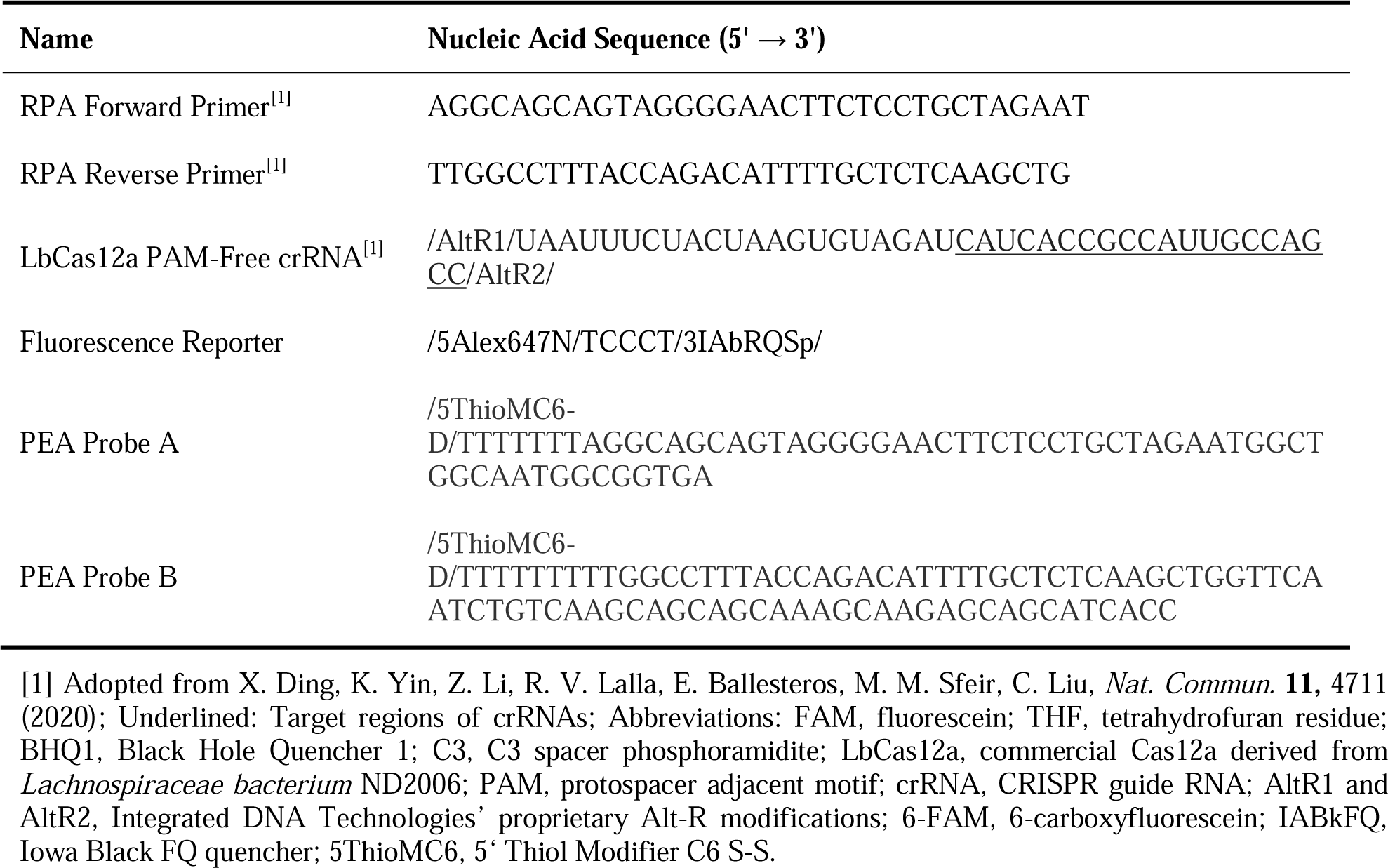
Sequences of RPA primers, fluorescence exo probe, Cas12a crRNAs, fluorescence reporter, and DNA activators.

## References

1. Chertow, D. S. Next-generation diagnostics with CRISPR. science 2018, 360, 381–382.

2. Li, Y.; Li, S.; Wang, J.; Liu, G. CRISPR/Cas systems towards next-generation biosensing. Trends Biotechnol. 2019, 37, 730–743.

3. Kaminski, M. M.; Abudayyeh, O. O.; Gootenberg, J. S.; Zhang, F.; Collins, J. J. CRISPR-based diagnostics. Nat. Biomed. Eng. 2021, 5, 643–656.

4. Kostyusheva, A.; Brezgin, S.; Babin, Y.; Vasilyeva, I.; Glebe, D.; Kostyushev, D.; Chulanov, V. CRISPR-Cas systems for diagnosing infectious diseases. Methods 2022, 203, 431–446.

5. Kumar, P.; Malik, Y. S.; Ganesh, B.; Rahangdale, S.; Saurabh, S.; Natesan, S.; Srivastava, A.; Sharun, K.; Yatoo, M. I.; Tiwari, R. CRISPR-Cas system: an approach with potentials for COVID-19 diagnosis and therapeutics. Frontiers in cellular and infection microbiology 2020, 10, 576875.

6. Hsieh, K.; Zhao, G.; Wang, T.-H. Applying biosensor development concepts to improve preamplification-free CRISPR/Cas12a-Dx. Analyst 2020, 145, 4880–4888.

7. Amiri-Dashatan, N.; Koushki, M.; Abbaszadeh, H.-A.; Rostami-Nejad, M.; Rezaei-Tavirani, M. Proteomics applications in health: biomarker and drug discovery and food industry. Iranian journal of pharmaceutical research: IJPR 2018, 17, 1523.

8. Borrebaeck, C. A. Precision diagnostics: moving towards protein biomarker signatures of clinical utility in cancer. Nature Reviews Cancer 2017, 17, 199–204.

9. Chan, H.-N.; Xu, D.; Ho, S.-L.; Wong, M. S.; Li, H.-W. Ultra-sensitive detection of protein biomarkers for diagnosis of Alzheimer’s disease. Chemical science 2017, 8, 4012–4018.

10. Zhou, S.; Lu, X.; Chen, C.; Sun, D. An immunoassay method for quantitative detection of proteins using single antibodies. Analytical biochemistry 2010, 400, 213–218.

11. Hartl, J.; Kurth, F.; Kappert, K.; Horst, D.; Mülleder, M.; Hartmann, G.; Ralser, M. Quantitative protein biomarker panels: a path to improved clinical practice through proteomics. EMBO Molecular Medicine 2023, 15, e16061.

12. Landegren, U.; Hammond, M. Cancer diagnostics based on plasma protein biomarkers: hard times but great expectations. Molecular oncology 2021, 15, 1715–1726.

13. Schiess, R.; Wollscheid, B.; Aebersold, R. Targeted proteomic strategy for clinical biomarker discovery. Molecular oncology 2009, 3, 33–44.

14. Basil, C. F.; Zhao, Y.; Zavaglia, K.; Jin, P.; Panelli, M. C.; Voiculescu, S.; Mandruzzato, S.; Lee, H. M.; Seliger, B.; Freedman, R. S. Common cancer biomarkers. Cancer research 2006, 66, 2953–2961.

15. Zangar, R. C.; Daly, D. S.; White, A. M. ELISA microarray technology as a high-throughput system for cancer biomarker validation. Expert review of proteomics 2006, 3, 37–44.

16. Barker, A. D.; Alba, M. M.; Mallick, P.; Agus, D. B.; Lee, J. S. An Inflection Point in Cancer Protein Biomarkers: What Was and What’s Next. Molecular & Cellular Proteomics 2023, 100569.

17. Zhao, T.; Hu, Y.; Zang, T.; Wang, Y. Identifying Protein Biomarkers in Blood for Alzheimer’s Disease. Frontiers in Cell and Developmental Biology 2020, 472.

18. Pérez, M.; Hernández, F.; Avila, J. Protein biomarkers for the diagnosis of Alzheimer’s disease at different stages of neurodegeneration. Int. J. Mol. Sci. 2020, 21, 6749.

19. May, A.; Wang, T. J. Biomarkers for cardiovascular disease: challenges and future directions. Trends in molecular medicine 2008, 14, 261–267.

20. Wong, Y.-K.; Tse, H.-F. Circulating biomarkers for cardiovascular disease risk prediction in patients with cardiovascular disease. Frontiers in Cardiovascular Medicine 2021, 8, 713191.

21. Yang, M.; Shi, K.; Liu, F.; Kang, W.; Lei, C.; Nie, Z. Coupling of proteolysis-triggered transcription and CRISPR-Cas12a for ultrasensitive protease detection. Science China Chemistry 2021, 64, 330–336.

22. Kim, H.; Lee, S.; Yoon, J.; Song, J.; Park, H. G. CRISPR/Cas12a collateral cleavage activity for simple and rapid detection of protein/small molecule interaction. Biosens. Bioelectron. 2021, 194, 113587.

23. Li, B.; Shao, Z.; Chen, Y. An exonuclease protection and CRISPR/Cas12a integrated biosensor for the turn-on detection of transcription factors in cancer cells. Anal. Chim. Acta 2021, 1165, 338478.

24. Chen, Q.; Tian, T.; Xiong, E.; Wang, P.; Zhou, X. CRISPR/Cas13a signal amplification linked immunosorbent assay for femtomolar protein detection. Anal. Chem. 2019, 92, 573–577.

25. Lee, I.; Kwon, S.-J.; Sorci, M.; Heeger, P. S.; Dordick, J. S. Highly sensitive immuno-CRISPR assay for CXCL9 detection. Anal. Chem. 2021, 93, 16528–16534.

26. Li, Y.; Mansour, H.; Watson, C. J.; Tang, Y.; MacNeil, A. J.; Li, F. Amplified detection of nucleic acids and proteins using an isothermal proximity CRISPR Cas12a assay. Chemical Science 2021, 12, 2133–2137.

27. Li, Y.; Deng, F.; Goldys, E. M. A simple and versatile CRISPR/Cas12a-based immunosensing platform: Towards attomolar level sensitivity for small protein diagnostics. Talanta 2022, 246, 123469.

28. Assarsson, E.; Lundberg, M.; Holmquist, G.; Björkesten, J.; Bucht Thorsen, S.; Ekman, D.; Eriksson, A.; Rennel Dickens, E.; Ohlsson, S.; Edfeldt, G. Homogenous 96-plex PEA immunoassay exhibiting high sensitivity, specificity, and excellent scalability. PloS one 2014, 9, e95192.

29. Fredriksson, S.; Gullberg, M.; Jarvius, J.; Olsson, C.; Pietras, K.; Gústafsdóttir, S. M.; Östman, A.; Landegren, U. Protein detection using proximity-dependent DNA ligation assays. Nature biotechnology 2002, 20, 473–477.

30. Lundberg, M.; Eriksson, A.; Tran, B.; Assarsson, E.; Fredriksson, S. Homogeneous antibody-based proximity extension assays provide sensitive and specific detection of low-abundant proteins in human blood. Nucleic acids research 2011, 39, e102–e102.

31. Carlyle, B. C.; Kitchen, R. R.; Mattingly, Z.; Celia, A. M.; Trombetta, B. A.; Das, S.; Hyman, B. T.; Kivisäkk, P.; Arnold, S. E. Technical Performance Evaluation of Olink Proximity Extension Assay for Blood-Based Biomarker Discovery in Longitudinal Studies of Alzheimer’s Disease. Frontiers in Neurology 2022, 13, 889647.

32. Thorsen, S. B.; Lundberg, M.; Villablanca, A.; Christensen, S. L. T.; Belling, K. C.; Nielsen, B. S.; Knowles, M.; Gee, N.; Nielsen, H. J.; Brünner, N. Detection of serological biomarkers by proximity extension assay for detection of colorectal neoplasias in symptomatic individuals. Journal of translational medicine 2013, 11, 1–13.

33. Zhang, P.; Hu, J.; Park, J. S.; Hsieh, K.; Chen, L.; Mao, A.; Wang, T.-H. Highly Sensitive Serum Protein Analysis Using Magnetic Bead-Based Proximity Extension Assay. Anal. Chem. 2022, 94, 12481–12489.

34. Shao, F.; Park, J. S.; Zhao, G.; Hsieh, K.; Wang, T.-H. Elucidating the Role of CRISPR/Cas in Single-Step Isothermal Nucleic Acid Amplification Testing Assays. Anal. Chem. 2023, 95, 3873–3882.

35. Kellner, M. J.; Koob, J. G.; Gootenberg, J. S.; Abudayyeh, O. O.; Zhang, F. SHERLOCK: nucleic acid detection with CRISPR nucleases. Nature protocols 2019, 14, 2986–3012.

36. Guanghui, T.; Gong, J.; Kan, L.; Zhang, X.; He, Y.; Pan, J.; Zhao, L.; Tian, J.; Lin, S.; Lu, Z. A general onepot-method for nucleic acid detection with CRISPR-Cas12a. 2020.

37. Ding, X.; Yin, K.; Li, Z.; Lalla, R. V.; Ballesteros, E.; Sfeir, M. M.; Liu, C. Ultrasensitive and visual detection of SARS-CoV-2 using all-in-one dual CRISPR-Cas12a assay. Nat. Commun. 2020, 11, 1–10.

38. Park, J. S.; Hsieh, K.; Chen, L.; Kaushik, A.; Trick, A. Y.; Wang, T. H. Digital CRISPR/Cas Assisted assay for rapid and sensitive detection of SARS CoV 2. Adv. Sci. 2021, 8, 2003564.

39. Chen, J. S.; Ma, E.; Harrington, L. B.; Da Costa, M.; Tian, X.; Palefsky, J. M.; Doudna, J. A. CRISPR-Cas12a target binding unleashes indiscriminate single-stranded DNase activity. Science 2018, 360, 436–439.

40. Arizti-Sanz, J.; Freije, C. A.; Stanton, A. C.; Petros, B. A.; Boehm, C. K.; Siddiqui, S.; Shaw, B. M.; Adams, G.; Kosoko-Thoroddsen, T.-S. F.; Kemball, M. E. Streamlined inactivation, amplification, and Cas13-based detection of SARS-CoV-2. Nat. Commun. 2020, 11, 1–9.

41. Feng, W.; Peng, H.; Xu, J.; Liu, Y.; Pabbaraju, K.; Tipples, G.; Joyce, M. A.; Saffran, H. A.; Tyrrell, D. L.; Babiuk, S. Integrating Reverse Transcription Recombinase Polymerase Amplification with CRISPR Technology for the One-Tube Assay of RNA. Analytical Chemistry 2021, 93, 12808–12816.

42. Ding, X.; Yin, K.; Li, Z.; Sfeir, M. M.; Liu, C. Sensitive quantitative detection of SARS-CoV-2 in clinical samples using digital warm-start CRISPR assay. Biosens. Bioelectron. 2021, 184, 113218.

43. Kaur, J.; Preethi, M.; Srivastava, R.; Borse, V. Role of IL-6 and IL-8 biomarkers for optical and electrochemical based point-of-care detection of oral cancer. Biosensors and Bioelectronics: X 2022, 11, 100212.

44. Bazzichetto, C.; Milella, M.; Zampiva, I.; Simionato, F.; Amoreo, C. A.; Buglioni, S.; Pacelli, C.; Le Pera, L.; Colombo, T.; Bria, E. Interleukin-8 in colorectal cancer: a systematic review and meta-analysis of its potential role as a prognostic biomarker. Biomedicines 2022, 10, 2631.

45. Principe, S.; Zapater-Latorre, E.; Arribas, L.; Garcia-Miragall, E.; Bagan, J. Salivary IL-8 as a putative predictive biomarker of radiotherapy response in head and neck cancer patients. Clinical Oral Investigations 2022, 1–12.

46. Gremese, E.; Tolusso, B.; Bruno, D.; Perniola, S.; Ferraccioli, G.; Alivernini, S. The forgotten key players in rheumatoid arthritis: IL-8 and IL-17–Unmet needs and therapeutic perspectives. Frontiers in Medicine 2023, 10, 956127.

47. Lemster, B.; Carroll, P.; Rilo, H.; Johnson, N.; Nikaein, A.; Thomson, A. IL-8/IL-8 receptor expression in psoriasis and the response to systemic tacrolimus (FK506) therapy. Clinical & Experimental Immunology 1995, 99, 148–154.

48. Reynolds, C. J.; Quigley, K.; Cheng, X.; Suresh, A.; Tahir, S.; Ahmed-Jushuf, F.; Nawab, K.; Choy, K.; Walker, S. A.; Mathie, S. A. Lung defense through IL-8 carries a cost of chronic lung remodeling and impaired function. American journal of respiratory cell and molecular biology 2018, 59, 557–571.

49. Apostolakis, S.; Vogiatzi, K.; Amanatidou, V.; Spandidos, D. A. Interleukin 8 and cardiovascular disease. Cardiovascular research 2009, 84, 353–360.

50. Dechkhajorn, W.; Maneerat, Y.; Prasongsukarn, K.; Kanchanaphum, P.; Kumsiri, R. Interleukin 8 in Hyperlipidemia and Coronary Heart Disease in Thai Patients Taking Statin Cholesterol Lowering Medication While Undergoing Coronary Artery Bypass Grafting Treatment. Scientifica 2020, 2020, 5843958.

51. Capogna, E.; Watne, L. O.; Sørensen, Ø.; Guichelaar, C. J.; Idland, A. V.; Halaas, N. B.; Blennow, K.; Zetterberg, H.; Walhovd, K. B.; Fjell, A. M. Associations of neuroinflammatory IL-6 and IL-8 with brain atrophy, memory decline, and core AD biomarkers–in cognitively unimpaired older adults. *Brain*, Behavior, and Immunity 2023, 113, 56–65.

52. Zhu, Y.; Chai, Y. L.; Hilal, S.; Ikram, M. K.; Venketasubramanian, N.; Wong, B.-S.; Chen, C. P.; Lai, M. K. Serum IL-8 is a marker of white-matter hyperintensities in patients with Alzheimer’s disease. *Alzheimer’s & Dementia: Diagnosis*, Assessment & Disease Monitoring 2017, 7, 41–47.

53. Heidt, B.; Siqueira, W. F.; Eersels, K.; Diliën, H.; van Grinsven, B.; Fujiwara, R. T.; Cleij, T. J. Point of care diagnostics in resource-limited settings: A review of the present and future of PoC in its most needed environment. Biosensors 2020, 10, 133.

54. Kumar, S.; Nehra, M.; Khurana, S.; Dilbaghi, N.; Kumar, V.; Kaushik, A.; Kim, K.-H. Aspects of point-of-care diagnostics for personalized health wellness. International journal of nanomedicine 2021, 383–402.

55. Piorino, F.; Patterson, A. T.; Styczynski, M. P. Low-cost, point-of-care biomarker quantification. Current opinion in biotechnology 2022, 76, 102738.

56. Park, J. S.; Akarapipad, P.; Chen, F.-E.; Shao, F.; Mostafa, H.; Hsieh, K.; Wang, T.-H. Digitized Kinetic Analysis Enhances Genotyping Capacity of CRISPR-Based Biosensing. ACS nano 2024, 18, 18058–18070.

57. Politza, A. J.; Nouri, R.; Guan, W. Digital CRISPR systems for the next generation of nucleic acid quantification. TrAC Trends in Analytical Chemistry 2023, 159, 116917.

58. Darmanis, S.; Nong, R. Y.; Hammond, M.; Gu, J.; Alderborn, A.; Vänelid, J.; Siegbahn, A.; Gustafsdottir, S.; Ericsson, O.; Landegren, U. Sensitive plasma protein analysis by microparticle-based proximity ligation assays. Molecular & cellular proteomics 2010, 9, 327–335.

59. Yelleswarapu, V.; Buser, J. R.; Haber, M.; Baron, J.; Inapuri, E.; Issadore, D. Mobile platform for rapid sub–picogram-per-milliliter, multiplexed, digital droplet detection of proteins. Proc. Natl. Acad. Sci. U.S.A. 2019, 116, 4489–4495.

60. Piepenburg, O.; Williams, C. H.; Stemple, D. L.; Armes, N. A. DNA detection using recombination proteins. PLoS biology 2006, 4, e204.

61. Notomi, T.; Okayama, H.; Masubuchi, H.; Yonekawa, T.; Watanabe, K.; Amino, N.; Hase, T. Loop-mediated isothermal amplification of DNA. Nucleic acids research 2000, 28, e63–e63.

62. Mori, Y.; Kitao, M.; Tomita, N.; Notomi, T. Real-time turbidimetry of LAMP reaction for quantifying template DNA. Journal of biochemical and biophysical methods 2004, 59, 145–157.

63. Banerji, U.; Judson, I.; Workman, P. The clinical applications of heat shock protein inhibitors in cancer-present and future. Current cancer drug targets 2003, 3, 385–390.

64. Rask-Andersen, M.; Zhang, J.; Fabbro, D.; Schiöth, H. B. Advances in kinase targeting: current clinical use and clinical trials. Trends in pharmacological sciences 2014, 35, 604–620.

65. Sandow, J. J.; Rainczuk, A.; Infusini, G.; Makanji, M.; Bilandzic, M.; Wilson, A. L.; Fairweather, N.; Stanton, P. G.; Garama, D.; Gough, D. Discovery and validation of novel protein biomarkers in ovarian cancer patient urine. PROTEOMICS–Clinical Applications 2018, 12, 1700135.

66. Bos, S.; Phillips, M.; Watts, G. F.; Verhoeven, A. J.; Sijbrands, E. J.; Ward, N. C. Novel protein biomarkers associated with coronary artery disease in statin-treated patients with familial hypercholesterolemia. Journal of clinical lipidology 2017, 11, 682–693.

67. Akinkuolie, A. O.; Buring, J. E.; Ridker, P. M.; Mora, S. A novel protein glycan biomarker and future cardiovascular disease events. Journal of the American Heart Association 2014, 3, e001221.

68. Lorenzo-Pouso, A. I.; Pérez-Sayáns, M.; Bravo, S. B.; López-Jornet, P.; García-Vence, M.; Alonso-Sampedro, M.; Carballo, J.; García-García, A. Protein based salivary profiles as novel biomarkers for oral diseases. Disease markers 2018, 2018, 6141845.

69. Yu, S.; Liu, Y.-P.; Liu, H.-L.; Li, J.; Xiang, Y.; Liu, Y.-H.; Jiao, S.-S.; Liu, L.; Wang, Y.; Fu, W. Serum protein-based profiles as novel biomarkers for the diagnosis of Alzheimer’s disease. Molecular Neurobiology 2018, 55, 3999–4008.

70. Hu, S.; Loo, J. A.; Wong, D. T. Human body fluid proteome analysis. Proteomics 2006, 6, 6326–6353.

71. Chen, F.-E.; Lee, P.-W.; Trick, A. Y.; Park, J. S.; Chen, L.; Shah, K.; Mostafa, H.; Carroll, K. C.; Hsieh, K.; Wang, T.-H. Point-of-care CRISPR-Cas-assisted SARS-CoV-2 detection in an automated and portable droplet magnetofluidic device. Biosens. Bioelectron. 2021, 190, 113390.

72. Zhang, P.; Chen, L.; Hu, J.; Trick, A. Y.; Chen, F.-E.; Hsieh, K.; Zhao, Y.; Coleman, B.; Kruczynski, K.; Pisanic II, T. R. Magnetofluidic immuno-PCR for point-of-care COVID-19 serological testing. Biosens. Bioelectron. 2022, 195, 113656.

73. Zhao, Y.; Chen, F.; Li, Q.; Wang, L.; Fan, C. Isothermal amplification of nucleic acids. Chem. Rev. 2015, 115, 12491–12545.

74. Craw, P.; Balachandran, W. Isothermal nucleic acid amplification technologies for point-of-care diagnostics: a critical review. Lab Chip 2012, 12, 2469–2486.

75. Ngo, H. T.; Akarapipad, P.; Lee, P.-W.; Park, J. S.; Chen, F.-E.; Trick, A. Y.; Wang, T.-H.; Hsieh, K. Rapid and portable quantification of HIV RNA via a smartphone-enabled digital CRISPR device and deep learning. Sensors and Actuators Reports 2024, 100212.

76. Shao, F.; Li, H.; Hsieh, K.; Zhang, P.; Li, S.; Wang, T.-H. Automated and miniaturized screening of antibiotic combinations via robotic-printed combinatorial droplet platform. Acta Pharmaceutica Sinica B 2023.

77. Zhang, P.; Kaushik, A. M.; Hsieh, K.; Li, S.; Lewis, S.; Mach, K. E.; Liao, J. C.; Carroll, K. C.; Wang, T. H. A Cascaded Droplet Microfluidic Platform Enables High Throughput Single Cell Antibiotic Susceptibility Testing at Scale. Small Methods 2022, 6, 2101254.

